# Development and first-in-human CAR T therapy against the pathognomonic MiT-fusion driven protein GPNMB

**DOI:** 10.1101/2025.02.26.24319604

**Authors:** Franz J. Zemp, Zackariah Breckenridge, Hyojin Song, Gurveer S. Gill, Hongrui Liu, Yeon Suh, Louisa Guignard, Joanna Pyczek, Cini John, Laura K. Mah, Jahanara Rajwani, Taye Louie, Kristofor K. Ellestad, Danyel Evseev, Madison Turk, Sacha Benaoudia, Jonathan Alex, Victor Naumenko, Varsha Thoppey Manoharan, Mobina Kazemi Mehrabadi, Harmony Aisling Smith, Geneece N.Y. Gilbert, Kiran Narta, Aaron Gilmour, Ted B. Verhey, Hayley M. Todesco, Katalin Osz, Bo-Young Ahn, Ana Bogossian, Colleen Anderson, Tarek A. Bismar, Daniel Y. C. Heng, Keith Lawson, Marston Lineham, Haley Pedersen, Paul Gordon, John B. McIntyre, John MacGregor, Kathy Brodeur-Robb, Lisa Difrancesco, Travis Ogilvie, Patrick Schöffski, Agnieszka Wozniak, Robert A. Holt, John Bell, Donna Senger, Victor Lewis, Michael Monument, Kyle Potts, Kiril Trpkov, Kevin Hay, Jennifer Quizi, Jennifer Chan, Ramy Saleh, Jan Willem Henning, Nicole Prokopishyn, A. Sorana Morrissy, Mona Shafey, Douglas J. Mahoney

## Abstract

CAR T therapy for solid tumors is limited by a lack of safe and uniformly expressed cell-surface targets. Here, we identify the MiT fusion-driven protein GPNMB as being highly, homogeneously, and stably expressed in primary and relapsed translocation-positive alveolar soft part sarcoma (ASPS) and renal cell carcinoma (tRCC). We developed a GPNMB-targeting CAR T therapy called GCAR1 that shows activity against patient-matched cells, organoids and xenograft models. First-in-human treatment of a patient with metastatic ASPS was well tolerated and generated stable disease until 6 months, with many non-target lesions resolving post-treatment. A polyclonal population of GCAR1 cells expanded in blood and were detectable until 6 months. Spatial transcriptomics revealed multiple immunosuppressive niches in proximity to T cells infiltrating a treatment-resistant lesion, and PDL1 blockade showed synergy with GCAR1 in a xenograft model. Our data provide clinical evidence for treating solid tumors with CAR T cells targeting a surface protein driven by an oncogenic gene fusion.

## INTRODUCTION

CAR T therapy represents a potentially curative treatment for cancer. Indeed, up to 50% of patients with CD19^+^ malignancies experience complete and durable remission after CAR T therapy.^1^ While activity against solid tumors has been more limited, notable instances of deep responses have been reported.^2–4^ Unfortunately, the identification of molecules selectively expressed on the surface of solid tumors has proven difficult, and examples of serious “on-target off-tumor” toxicities have been observed.^5^ In addition, solid tumors are often heterogeneous, harboring populations of CAR target-negative cells that create a reservoir of treatment resistance.^3,6^ The discovery of surface proteins that are highly, uniformly and stably expressed on cancer cells, with minimal or manageable expression on normal tissues, remains a formidable challenge for the development of CAR T therapies for solid malignancies.

Approximately 30% of soft-tissue sarcomas (STS), as well as subtypes of carcinomas and leukemias, are molecularly characterized by chromosomal translocations that create novel oncogenes.^7^ Many are gene fusions that encode aberrantly-regulated transcription factors capable of driving cellular transformation, often in the absence of other genetic abnormalities.^8^ For example, the Microphthalmia-associated transcription factor (MiT) family, such as TFE3, are frequent 3’ partners in pathognomonic gene fusions driving alveolar soft-part sarcoma (ASPS), translocation RCC (tRCC), perivascular epithelioid cell neoplasms (PEComa) and some epithelioid hemangiosarcomas (EHE).^9^ These cancers often exhibit dependence on their gene fusions and subsequently dysregulated transcriptomes, leading to a phenomenon known as oncogene addiction.^10^ However, with a few notable exceptions such as BCR-ABL, oncogenic gene fusions and their aberrant transcriptomes have proven difficult to target.

In this study, we tested a hypothesis that MiT fusion-driven neoplasms express cell-surface proteins that could be effectively targeted with CAR T therapy, because those proteins are 1.) driven by the gene fusion, and thus aberrantly expressed to high, uniform and stable levels from the cancer, and 2.) important for disease biology. Following an unbiased proteogenomic surfaceome screen in ASPS, we developed a 2nd generation CAR T therapy (GCAR1) against our top candidate, glycoprotein non-metastatic B (GPNMB), a type Ia transmembrane protein directly regulated by MiT fusions and known to promote metastasis and cancer immune evasion in multiple cancer types.^11–13^ We report that GCAR1 is well tolerated and shows anticancer activity in patient-matched preclinical models of MiT fusion-derived cancers and a first-in-human study of a patient with relapsed/refractory, metastatic ASPS. As a precision immunotherapeutic, GCAR1 may have widespread applications against numerous cancer types derived from MiT gene fusions or those harboring oncogenic drivers of MiT activity. More broadly, our data provide support for targeting surface proteins driven by oncogenic gene fusions with CAR T therapeutics.

### GPNMB is highly expressed from MiT fusion-driven tumors

To identify potential CAR T cell targets in MiT fusion-driven tumors, we developed a computational pipeline for predicting highly expressed cell surface proteins using tumour gene expression data generated from a cohort of ASPS patients. Our analyses revealed eleven candidate proteins (*Supplementary Fig. 1*), with GPNMB, shown previously to be a direct transcriptional target of MiT fusion proteins,^14,15^ being the most significantly upregulated. Subsequent analyses of additional ASPS patient cohorts found that GPNMB expression remains elevated post-treatment and in metastases (**Fig. 1a**), with levels notably higher than in other pediatric, adolescent and young adult tumors not driven by MiT fusion genes (**Fig. 1b**). Protein validation by immunohistochemistry (IHC) revealed high and homogeneous GPNMB staining (H-score ≥200) in 82.3% of ASPS samples on a tissue microarray (TMA) comprising 62 primary or metastatic lesions^16^ (**Fig. 1c**). Further, IHC analysis of eight primary and recurrent lesions resected from six organs over a six-year period from a single ASPS patient (Patient ASPS-1) demonstrated spatiotemporal stability of GPNMB expression (**Fig. 1d,e)**. This was also observed in the matched primary and metastasis samples on the TMA (**Fig. 1f,g**). Similarly high and homogenous GPNMB expression (H-score ≥200 in 80% of samples tested) was observed in a cohort of patients with MiT fusion-driven RCC (**Fig 1h,i**), consistent with a recent report.^17^

**Figure 1:**
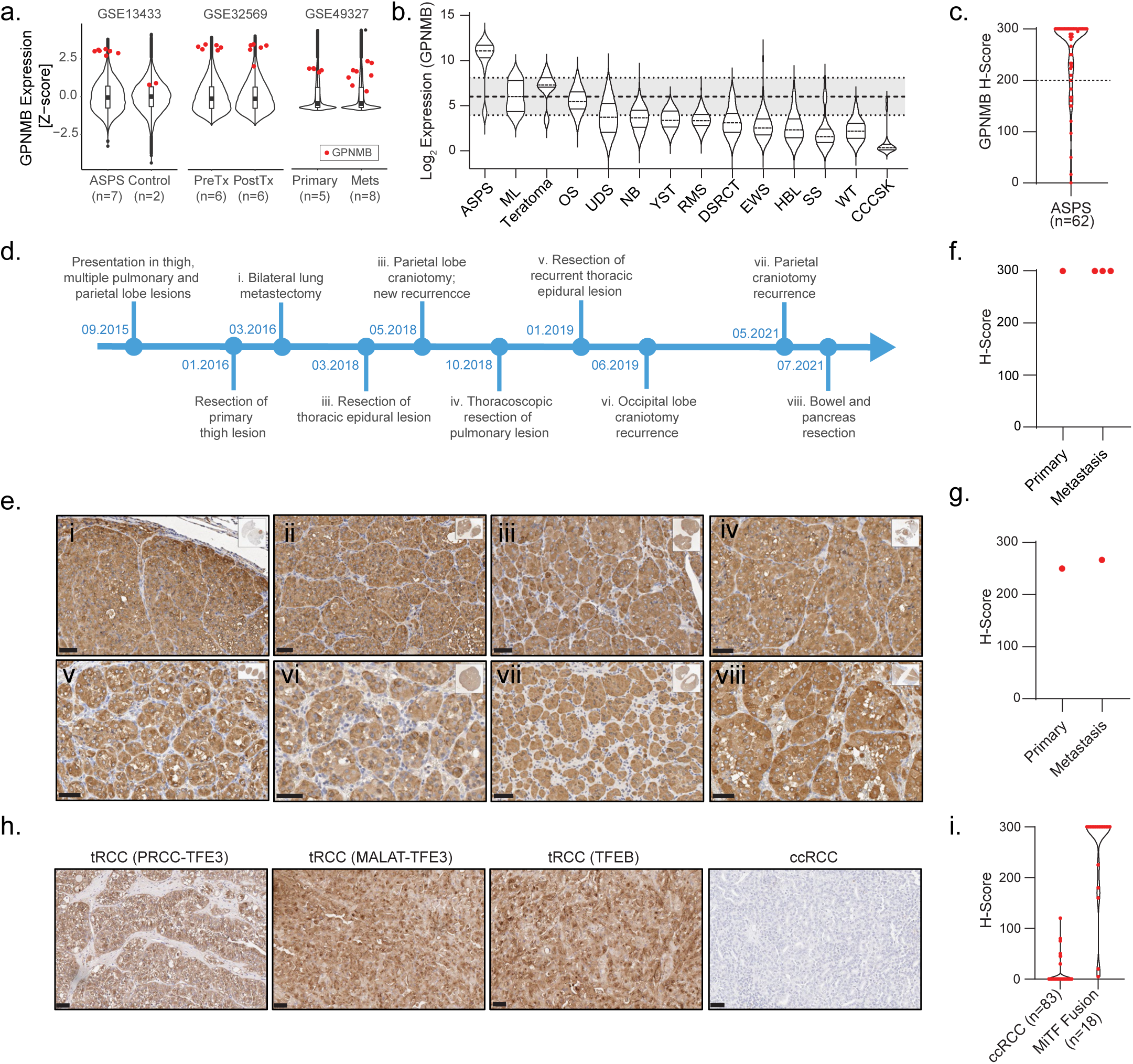
GPNMB is highly expressed in MiT fusion-driven tumors. **a,** Gene expression levels of GPNMB (red) across three publicly available ASPS databases compared to normal tissue (GSE13433), post systemic therapy (GSE32569) and in primary vs. metastatic lesions (GSE49327). Expression data is presented as Z-scores. Violin plots include all genes in all patients per category; box plots indicate median values (black line), the 25-75 percentile (box), and lines span the data range. **b,** Violin plot of GPNMB expression in whole-transcriptome sequencing analysis of 657 pediatric/AYA extracranial solid cancer samples. Dashed line is median, solid line is quartiles. Grey box is the expression of 37-housekeeping genes in each sample. Dashed line within box is median house-keeping gene expression; Dotted line is one standard deviation of the mean. **c,** Tissue microarray of 62 ASPS patient tumor samples stained and scored for GPNMB. **d,** Surgical time course of patient ASPS-1 (no tissue obtained from stereotactic radiosurgery of right parietal lobe lesion present at diagnosis, 02-2016). **e,** GPNMB protein levels in each of these resections, roman numerals correspond to dates on time course. Scale bars = 50μm. **f-g**, GPNMB expression in matched patient primary and metastasis samples. **h-i**, Immunohistochemical staining GPNMB and scoring of different MiT fusion positive renal cell carcinoma cases (tRCC) versus clear cell renal cell carcinoma (ccRCC). Scale bars = 50μm.

GPNMB is a type Ia transmembrane glycoprotein with diverse functions and expression across normal tissues, but can be upregulated by inflammation and in cancer.^11,13^ Although steady-state GPNMB expression has been reported in the skin, bone, and subsets of myeloid-derived cells,^13^ our interrogation of GPNMB protein levels in publicly available mass-spectrometry databases revealed low or undetectable expression in all normal tissues compared to a panel of housekeeping genes, and comparable to two common CAR targets in solid tumors (B7-H3 and HER2; *Supplementary Fig. 2*). We further examined GPNMB expression by histology in 34 normal tissues across two normal tissue microarrays. Moderate to marked (2-3+) GPNMB expression was noted in the placental trophoblasts and macrophages in the tonsil, lung and liver (*Supplementary Table 1*).

### CAR T cells targeting GPNMB are efficacious in preclinical models of MiT fusion-driven cancer

Considering that GPNMB is highly, stably and homogeneously expressed from MiT-driven tumours, and its expression in normal tissue is relatively low, we developed a collection of second-generation (41BBζ) GPNMB-targeting CARs (**Extended Data Fig. 1a**) and tested them for activity when engineered into primary human T cells. The most active (G1), designated GCAR1, incorporates the fully-humanized scFv within the CR011 antibody clone.^18^ It showed robust CAR surface expression, potent cytotoxicity and high secretion of IL-2 and IFNγ when incubated with an ASPS cell line cultured from a patient-derived xenograft (PDX) developed from one of Patient ASPS-1’s lung metastases (**Extended Data Fig. 1b-g**; *Supplementary Table 2*). These findings were validated using T cells harvested from two additional healthy donors (**Extended Data Fig. 1h-i**).

We then assessed GCAR1 efficacy against a xenograft model of ASPS, established using the same patient cell line. Consistent with the patient’s primary tumour sample, this xenograft retained the type II ASPSCR1-TFE3 fusion, exhibited nuclear localization of TFE3, high GPNMB expression, and histological characteristics of ASPS (*Supplementary Fig. 3*). Tumor reduction was observed 4 days post-treatment with a single dose of 5e6 or 1e6 GCAR1 cells, with complete tumor elimination achieved between 7 and 14 days (**Extended Data Fig. 2a-c**). A dose of 1e5 GCAR1 cells elicited a partial tumor response, with maximal reduction at D21 and a modest survival advantage. Anti-tumor activity correlated with GCAR1 expansion in blood, peaking at 1162- and 463-fold increase over CD19 CARs on D14 for the 5e6 and 1e6 dose, respectively (**Extended Data Fig. 2d**; *Supplementary Figure 4a*). CAR expression was transiently decreased in circulating CAR T cells (**Extended Data Fig. 2e**), which we suspect was caused by activation-induced CAR internalization,^19^ as EGFP expression remained stable. GCAR1 cells accumulated within the xenograft microenvironment (**Extended Data Fig. 2f-g**) and were more stationary than CD19-CAR T cells (**Extended Data Fig. 2h**), consistent with target engagement.^20,21^

ASPS has the highest incidence of brain metastasis among sarcomas (11-19%),^22^ which is a significant cause of morbidity and mortality. Intravascular dosing with 1e6 or 1e5 GCAR1 cells completely eradicated ASPS tumors grown in the brain (**Extended Data Fig. 3a-d**). Spinal metastases were also resolved with GCAR1. Tumor regression correlated with CAR T cell expansion in blood, though expansion was less than in the primary tumor model, peaking at a 16.5-fold increase over non-targeting CD19 CARs at 14 days post-treatment (**Extended Data Fig. 3e**). While 1e6 GCAR1 cells induced rapid weight loss, it resolved within 7 days (**Extended Data Fig. 3f**) and neuropathological assessment showed no residual tumor or neurological damage. We suspect that this transient toxicity, observed only at the highest dose, was due to rapid CAR T cell expansion and subsequent inflammation in the intracranial space, and not on-target off-tumor toxicity, as the GCAR1 scFv does not target mouse GPNMB (*Supplementary Figure 5)*.^18^

### Patient-derived GCAR1 is active against matched preclinical cancer models

As healthy donor-derived GCAR1 was efficacious in preclinical models and there are few treatment options available for patients with metastatic ASPS, we conducted late preclinical studies to support clinical translation of GCAR1 for ASPS and other MiT-driven cancers. To begin, we sought to determine whether GCAR1 could be successfully engineered from patient T cells to target a model of their own disease (**Fig. 2a**). As hoped, GCAR1 engineered from Patient ASPS-1 T cells induced cytotoxicity and secreted high-level cytokines when co-cultured with her ASPS cells *in vitro* (**Fig. 2b-c**). They also generated rapid tumor regression of her xenograft, with profound GCAR1 expansion in blood post-treatment (**Fig. 2d-f**). To validate these findings, we engineered GCAR1 from T cells freshly isolated from a second ASPS patient (Patient ASPS-2; *Supplementary Table 2, Supplementary Figure 6a*) and co-cultured them with explants of his primary tumor, a model system recently shown to correlate with CAR T cell efficacy in patients.^41^ These cells infiltrated and expanded in the explants (**Fig. 2g-h**) and the culture media accumulated molecules indicative of T cell activation, such as Granzyme B and IFNγ, amongst others (**Fig. 2i**, *Supplementary Figure 6b*).

**Figure 2:**
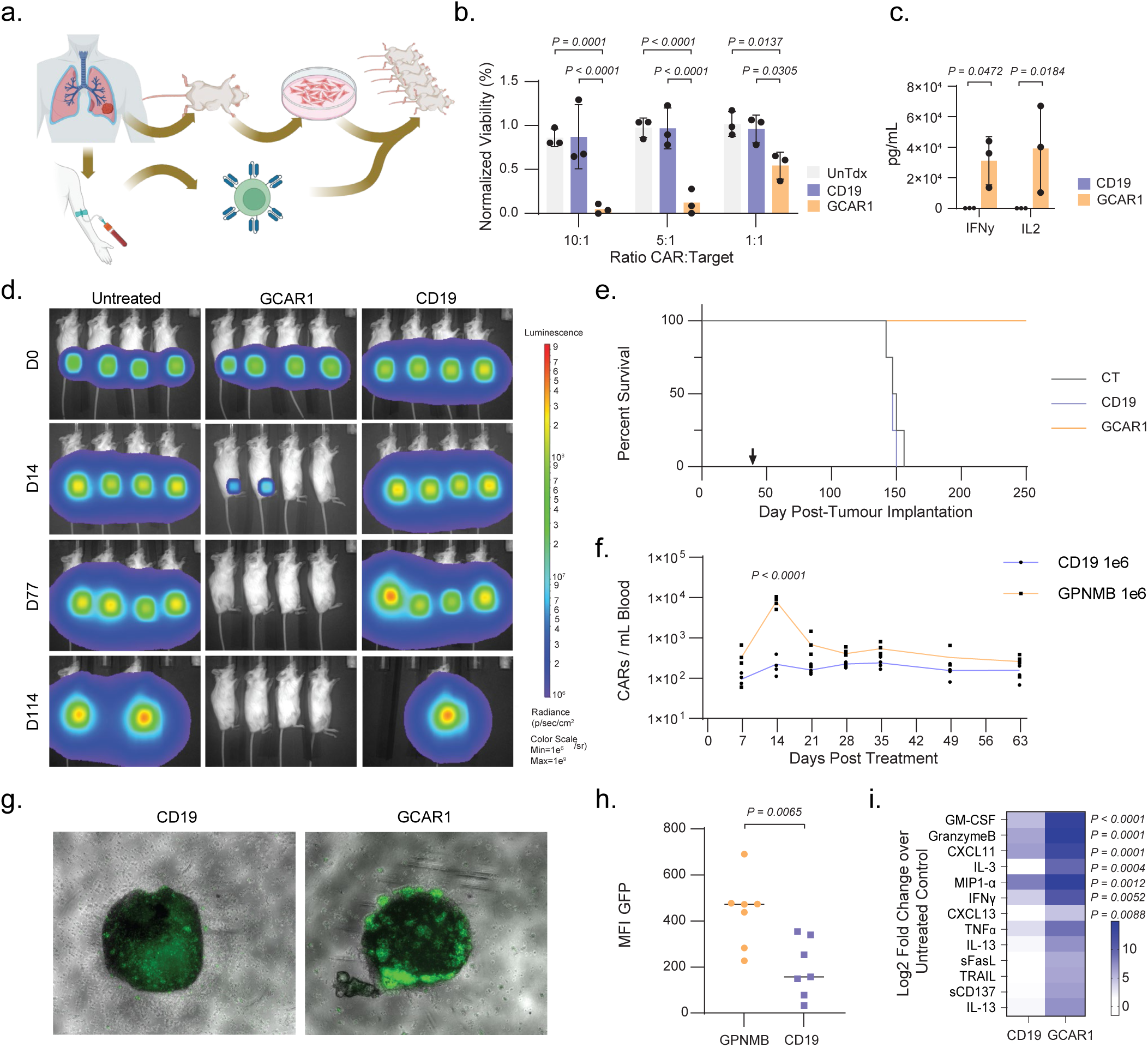
Patient-derived GPNMB-targeting CAR T cells are efficacious against matched preclinical cancer models. **a,** Schematic representation of model development matched with GCAR1 treatment for Patient ASPS-1. **b,** Cytotoxic activity of Patient ASPS-1 GCAR1 (orange) cells against her ASPS cell line, compared to a non-targeting CD19 CAR (blue) or non-transduced cells (grey). Blood was drawn and CAR T cells generated on three separate occasions (n=3). Bars are means with standard deviation. 2-way ANOVA, Tukey’s multiple comparisons test. **c,** Concentration of IFNγ and IL-2 in media harvested 24 hours after 1:1 co-culture of Patient ASPS-1 GCAR1 cells (orange) or CD19 (blue) CAR T cells with her ASPS cells. Bars are means with standard deviation. Unpaired T-test. **d,** Representative bioluminescent images from mice bearing intramuscular luciferase-expressing Patient ASPS-1 xenografts treated intravenously with 1e6 GCAR1 (n=4), CD19 CAR T (n=4) or untreated (n=4), manufactured from fresh Patient ASPS-1 PBMCs. **e,** Kaplan-Meier survival curve of mice from (d). **f,** Flow cytometry for GCAR1 cells in mice from (d). 2-way ANOVA with Šidak’s multiple comparisons test. **g,** Representative microscopy images of tumour explants from Patient ASPS-2 treated for 48 hours with 1e4 GFP^+^ CAR T cells manufactured from his PBMCs. **h,** Quantification of mean GFP intensity within the explant area. Line indicates median of all samples. n=7, unpaired T-test. **i,** Heatmap representation of cytokine concentrations in supernatants from (g-h). Heatmap represents the mean Log2-fold change over untreated control (n=3). 2-way ANOVA with Šidak’s multiple comparisons test.

### GCAR1 is safe and active in a first in-human study

Next, we engineered a “clinical lentiviral vector” by incorporating the scFv, hinge and MYC domains of our preclinical vector into a 2nd generation 41BBζ backbone currently in clinical trial (NCT03765177). In cell culture studies, GCAR1 engineered from the clinical vector performed comparably to that engineered from the preclinical vector (*Supplementary Figure 7a,b*). A batch of cGMP lentivirus was then produced and a GCAR1 manufacturing process developed on the CliniMACS Prodigy®. A series of validation runs yielded high-quality GCAR1 that met all predetermined release specifications and demonstrated robust activity against Patient ASPS-1 cells and xenograft model (*Supplementary Figure 7c-i*). The product also showed potent activity against tRCC cell lines and xenograft models expressing high levels of GPNMB (*Supplementary Figure 8*; **Extended Data Fig. 4**).

We then conducted a single patient study in a female in their 30’s with rapidly progressive ASPS (Patient ASPS-3; *Supplementary Table 2*), presenting prior to study with innumerable pulmonary metastasis that were not responding to atezolizumab (Patient History in **Extended Data Fig. 5a**; fusion status in *Supplementary Figure 9a-b*). The study’s primary objectives were to determine the feasibility, safety and efficacy of systemically delivered GCAR1, while secondarily assessing GCAR1 pharmacokinetics, phenotype, function, and the host’s response to treatment. GPNMB was highly expressed in her primary tumor (*Supplementary Figure 9c*) and GCAR1 manufacturing from 1e8 CD4/8^+^ cells enriched from fresh apheresis product resulted in 2.7e9 viable GCAR1 cells that were potent and met all release criteria (*Supplementary Figure 9d-h*). At study enrollment, the patient’s ECOG was 1 and her clinical symptoms were cough and mild shortness of breath on exertion. The study protocol was approved by Health Canada and the Health Research Ethics Board of Alberta.

ASPS Patient-3 received standard lymphodepleting chemotherapy with fludarabine and cyclophosphamide and a single dose of cryopreserved GCAR1, delivered intravenously at 1e6 CAR+ cells/kg (**Extended Data Fig. 5a**). Safety was evaluated using CTCAE grading of adverse event. Following GCAR1 therapy, the patient developed neutropenia, attributed to the lymphodepleting chemotherapy, that was uncomplicated and treated with a single dose of filgrastim on D13. She had mildly elevated transaminases (ALT/AST), C-reactive protein (CRP) and lactate dehydrogenase (LDH), which peaked between D13-16 and resolved without intervention within one week (**Fig. 3a**). On D14 she developed a Grade 1 urticarial rash on her upper limbs and abdomen (∼10% total BSA; **Fig. 3b**) that was resolved with topical betamethasone within 48 hours. She did not develop CRS or ICANS and her blood levels of CRS-associated inflammatory markers were only mildly elevated post-therapy (**Fig. 3c**). However, her blood levels of numerous cytokines and chemokines associated with cytotoxic T cell activity rose significantly after treatment, peaking on D14-D21 (*Supplementary Figure 10*).

**Figure 3:**
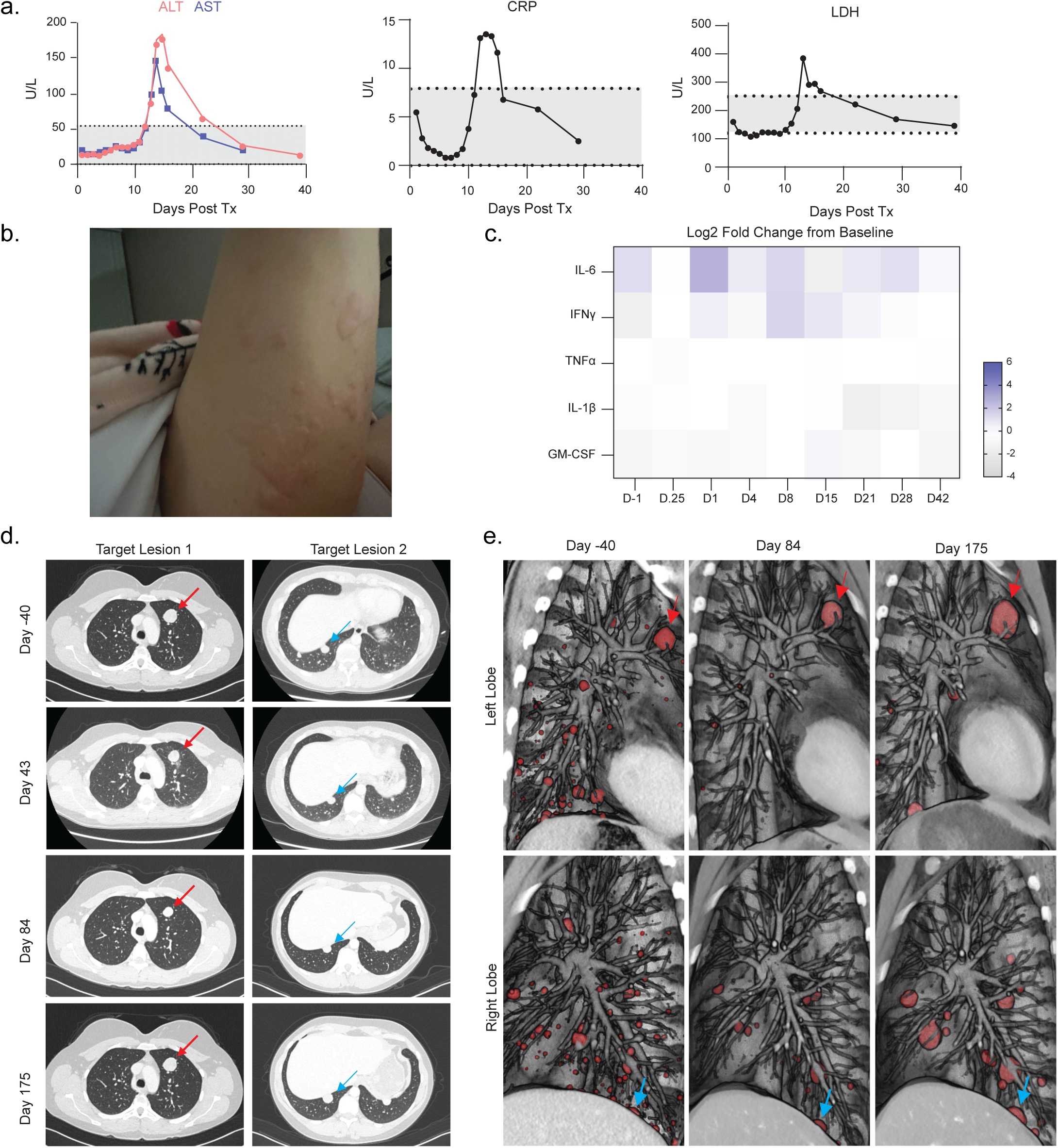
GCAR1 is safe and active in a first in-human single patient study. **a,** Blood concentration of C reactive protein (CRP), alanine transaminase (ALT), aspartate aminotransferase (AST) and lactate dehydrogenase (LDH) post-GCAR1 treatment. **b,** Photo of grade 1 urticarial rash on the patient’s arm 14 days post-GCAR1 treatment. **c,** Heatmap representation of concentration of plasma analytes associated with CRS post-GCAR1 therapy. Values represent Log2 transformed fold change over baseline measurements. Baseline was determined as the mean from three pretreatment visits. **d,** Single slice CT images of target lesions pre-GCAR1 pre-treatment (Day-40) and 43-, 84- and 176-days post-treatment. Red arrow indicates target lesion 1, Blue arrow indicates target lesion 2. **e,** Volumetric reconstruction of CT images pre- and post-GCAR1 therapy. Tumor lesions are pseudo-coloured in red. Red arrow indicates target lesion 1, Blue arrow indicates target lesion 2.

Treatment response was evaluated using RECIST 1.1.^23^ At baseline, the two largest lung lesions were designated target lesions, while the remaining innumerable lung lesions were considered non-target lesions. CT scans at 43- and 84-days post therapy showed that the target lesions remained stable in size (**Fig. 3d**). However, the non-target lesions were noticeably impacted by GCAR1, with near complete resolution of many diffuse pulmonary nodules seen at baseline **(Fig. 3e; Extended Data Fig. 5b**). Overall, the patient was deemed by independent radiology assessment to have experienced stable disease on D43 and 84. Importantly, her clinical respiratory symptoms were completely resolved at these timepoints. Unfortunately, CT imaging at 6 months post therapy revealed enlargement of both target lesions and some of the remaining non-target lesions. The patient is now experiencing progressive disease.

### GCAR1 undergoes polyclonal T cell expansion and evolution in blood

The patient’s absolute lymphocyte count (ALC) remained below normal range from D1-13 but rose dramatically into normal range from D14 onwards, reaching a peak at D16 (**Fig. 4a**). This coincided with peak GCAR1 expansion in blood as measured by droplet digital PCR (**Fig. 4b**) and flow cytometry (**Fig. 4c**). To generate deeper insight into these circulating GCAR1 cells, we characterized peripheral blood samples serially collected post-treatment at D08, 15, 22, 28, and 42 and compared them to the T cell-enriched apheresis product (Enriched) and the post-expansion GCAR1 harvest from the CliniMACS (Harvest). Based on single cell-level gene expression (GEX) data, including expression of the CAR construct, and matched profiling of the TCR repertoire (VDJ), we observed peak GCAR1^+^ cell expansion in blood at D15, consistent with other measurements of expansion (**Fig.4a-e**). Semi-automated cell type annotation^24^ of 47 GEX cell clusters identified 21 distinct cell types and their subclasses, including multiple T cell subsets (**Fig 4e-f; Extended Data Fig. 6a-d**, *Supplementary Figure 11*; *Supplementary Table 3*). Transcriptional T cell states shifted dramatically during GCAR1 production, from a CD4:CD8 ratio in the Enriched sample of 2:1, towards a CD4-dominant composition in the Harvest (**Fig 4h; Extended Data Fig 6e**). Intriguingly, expanded effector GCAR1^+^ cells predominant in circulation were mostly CD8^+^, suggesting that these cells expanded from a small starting population in the Harvest (**Fig 4 g-h; Extended Data Fig. 6e**). In particular, CD8^+^ effector (CD8e), effector memory (CD8em), and effector memory terminally differentiated (CD8em_td) cells comprised the vast majority of GCAR1^+^ cells at D08 (CD8e), D15 (CD8em; CD8em_td), and D22-28 samples (CD8em) (**Fig. 4g**), profiles also supported by flow cytometry (**Extended Data Fig. 6d**; *Supplementary Figure 4b*).

**Figure 4:**
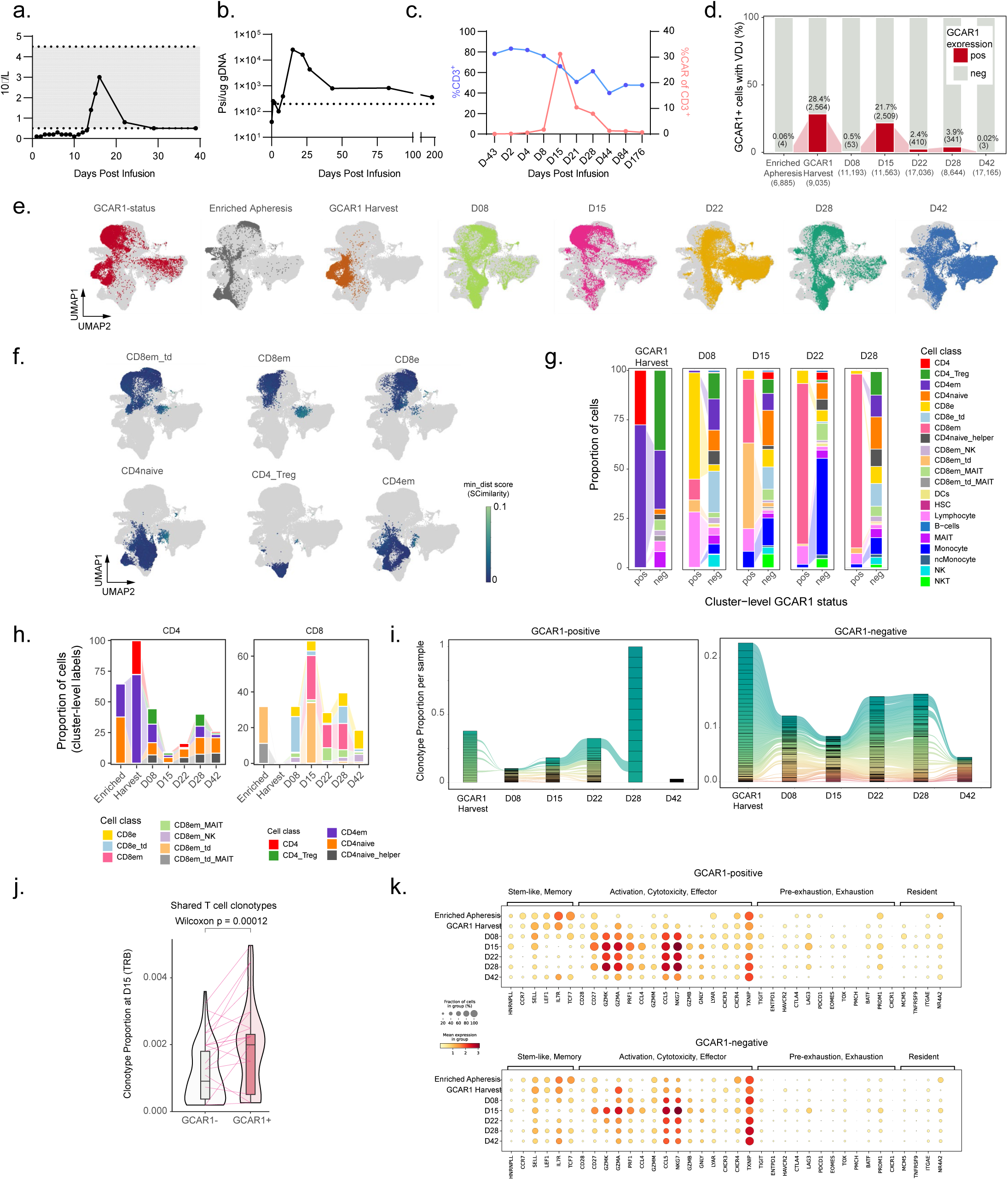
GCAR1 undergoes polyclonal T cell expansion and evolution in blood. **a,** Absolute lymphocyte count in blood post-GCAR1 treatment. **b,** CAR^+^ cells in blood post-GCAR1 treatment by droplet digital PCR. **c,** Total T cells (%CD3^+^ blue) and GCAR1^+^ cells (%CAR of CD3^+^ red) by flow cytometry. **d,** Stacked barplots of the proportion of GCAR1+ (red) and GCAR1- (grey) cells per sample. The total number of cells is listed under each sample label, the percent and number of GCAR1+ cells are annotated above the red bars. **e**, Uniform Manifold Approximation and Projection (UMAP) plots of cells from all samples. Subpanels highlight cells that are GCAR1^+^ (left panel) or belong to each of 7 longitudinal samples profiled. **f**, UMAP plots show cell-level annotations predicted by SCimilarity for a subset of cell types. Colour denotes the confidence of each label; 0 corresponds to maximum confidence (min_dist; see Methods). **g**, Paired stacked barplots of the proportion of cells in each sample that correspond to major cell classes, split by GCAR1 status. The Enriched Apheresis and D42 are not included as they do not have any or very few GCAR1^+^ cells. **h**, Stacked barplots of the proportion of cells across CD4 and CD8 subtypes. **i**, Alluvial plots of prevalent VDJ clonotypes from Harvest to D42 (6 samples; Apheresis excluded), are presented separately for GCAR1^+^ and GCAR1^-^ cells, using the union of the top 30 clonotypes per sample. Clonotypes were defined based on unique TRA and TRB nucleotide sequences and VDJC gene usage. **j**, Violin plot of GCAR1^+/-^ shared clonotypes (n=18; 11.8%), showing a significant increase in frequency within the GCAR1+ fraction. P-value is calculated using a Wilcoxon rank-sum test. Violin plots include all values; box plots indicate median values (black line), and the 25-75 percentile (box); lines connect clonotype frequencies between the two groups. **k**, Dot plot of CAR-T immunophenotyping marker gene expression by sample, segregated by GCAR1 status. Dots are scaled and coloured according to the fraction of cells and mean gene expression in each group, respectively.

TCR repertoire analysis demonstrated that GCAR1^+^ expansion was polyclonal, with immunodominant (top 30) clonotypes arising at D08 continuing to occupy a majority of the clonal space at D15 and D22 (**Fig. 4i**). Given the dramatic shift from CD4^+^ to CD8^+^ states between the Harvest and subsequent effector cells, most expanded CD8^+^ GCAR1^+^ clonotypes were not detectable in the Harvest. In contrast, the GCAR1^-^ fraction remained stable between D08-D42, and was broadly representative of the prevalent populations detected in the Harvest (**Fig. 4i**). A subset of GCAR1^+^ clonotypes (∼12%) were also detected in the GCAR1^-^ fraction. After infusion, the GCAR1^+^ cells in many of the shared clonotypes expanded to a significantly higher extent than the corresponding GCAR1^-^ cells (**Fig. 4j**).

GCAR1^+^ cells were strongly positive for T cell activation markers including *CD27*, *GZMA*, *GZMK*, *PRF1*, *CCL5*, and *NKG7* (**Fig 4k**), with activation evident at D08, peaking at D15 and strongly sustained to D28. By D42, the GCAR1^+^ cell population fell to the limit of detection (**Fig 4c-d**), in line with the reduction of metastatic disease burden (**Fig. 3d**). Exhaustion markers, most notably *LAG3*, *HAVCR2* (*TIM3*), and *TIGIT*, were also elevated at D15 (**Fig. 4k**). Flow cytometry confirmed protein-level upregulation of TIGIT and HAVCR2 (TIM3) while additionally identifying upregulation of PD1 (**Extended Data Fig. 6f**).

We noted a similar but less pronounced activation profile in the GCAR1-cell fraction at D15 (**Fig. 4k**), indicating the potential engagement of endogenous tumor-reactive T cells. Given that the ASPSCR1-TFE3 fusion is present in every cell, we speculated it may generate neoantigens capable of driving immune recognition. Considering both type I and type II fusions in patients ASPS-1 to -3 and an additional biobanked case (ASPS-4; *Supplementary Table 2*), we conducted an *in-silico* evaluation to determine whether fusion-derived neopeptides are presentable on MHC class-I. This revealed three type I (ERLPVSGNL, RERLPVSGNL and RERLPVSGNLL) and two type II (RIDDVIDEI and RIDDVIDEII) fusion neopeptides predicted by multiple algorithms to bind at least one MHC-I allele in the ASPS patients studied (**Extended Data Fig. 6g-j**; *Supplementary Tables 2,4*), or to a common MHC-I allele (>1% of the general population), extending observations made in a previous study.^25^ Importantly, the GCAR1-treated Patient ASPS-3 was predicted to present type I fusion neopeptides on C*07:02, providing a potential mechanism underlying the activation of endogenous GCAR1^-^ T cell responses in her tumour post-treatment.

### A GCAR1-refractory lung lesion harbours spatially distinct T cell inhibitory niches

Antigen-loss and immunosuppression in the TME are common mechanisms of relapse after CAR T therapy.^6,26^ IHC of a treatment-refractory lesion biopsied 35 days post-treatment demonstrated high-level GPNMB, ruling out antigen loss as the source of resistance (**Fig. 5a**). To investigate potential immunosuppressive mechanisms, we employed the 10X genomics Visium HD platform to generate transcriptome-wide gene expression profiles. GCAR1-specific probes were also spiked into the library. Using the data generated, we assessed the cellular composition of two D35 needle core biopsies sampled from a single GCAR1-refractory lesion, and from the patient’s primary tumor resection at diagnosis. These analyses confirmed high-level GPNMB expression was retained in the biopsy (**Fig 5b**). They also showed that while T cells were abundant in the biopsy **(Fig 5c,d)**, very few were GCAR1 positive, an observation confirmed by IHC (**Fig. 5a**).

**Figure 5:**
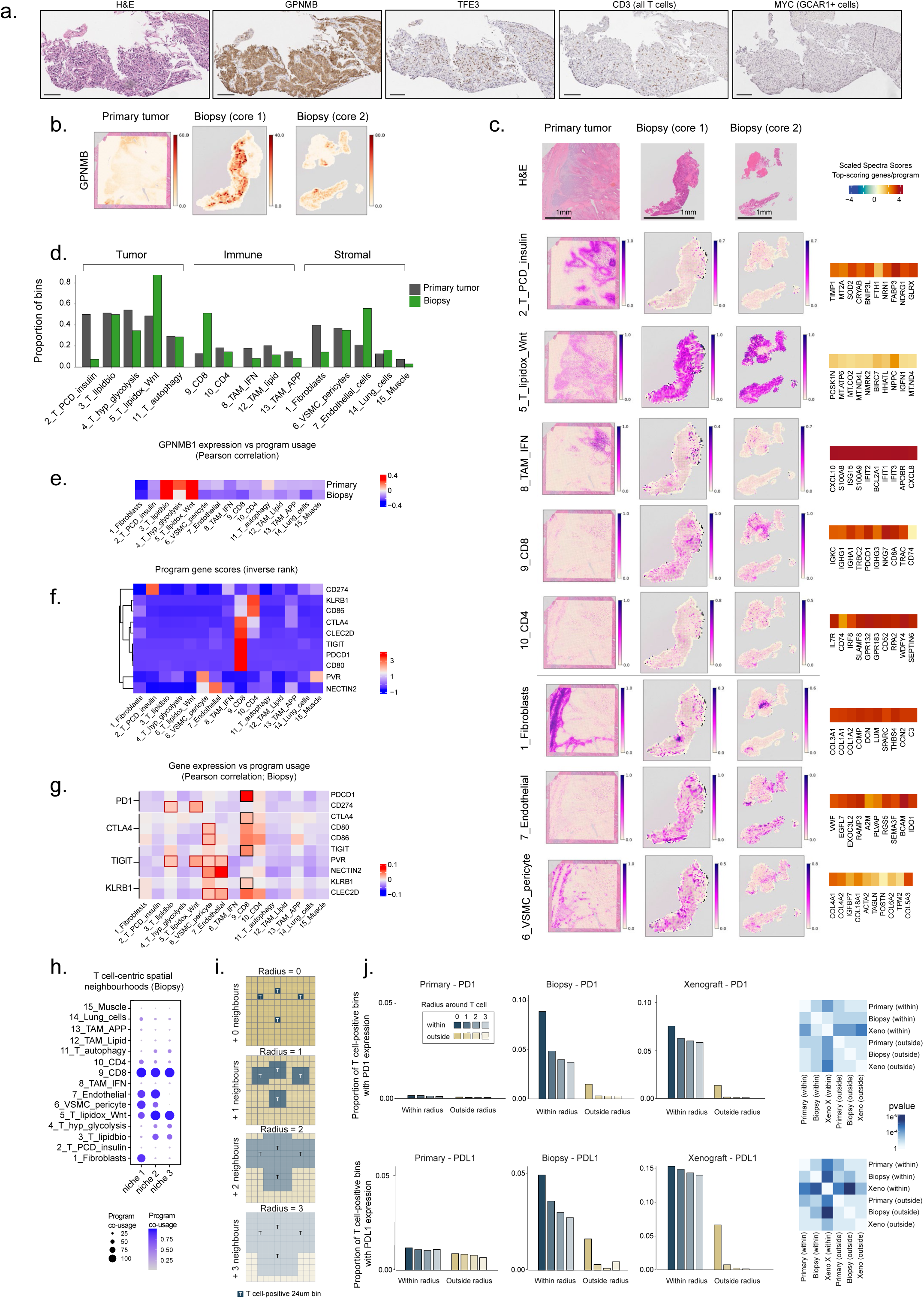
A GCAR1-refractory lung lesion harbours distinct T cell inhibitory niches. **a**, Hematoxylin and eosin (H&E; left panel) of lung tumour biopsy sample taken 35 days post-GCAR1 treatment, with (remaining panels) IHC staining for the CAR target GPNMB; TFE3 to mark tumor cells; CD3 to identify all T cells; MYC to identify GCAR1 cells. Scale bars = 100 μm. **b**, Spatial plot of GPNMB gene expression across three samples profiled with Visium HD, including the ASPS-3 patient primary tumor and 2 research core biopsies. **c**, H&E plots of the spatially profiled samples in (b), with spatial plots of gene expression program usage values, and top 10 scoring genes per program (heatmaps). **d**, Barplots of the proportion of 24 μm bins per sample that have a minimum usage of each program (usage >0.05). Primary (grey); biopsy (green). Biopsy samples are merged together. **e**, Pearson correlation between spatial GPNMB expression in each sample type (rows) and spatial program usage (columns). **f**, Heatmap plot of the inverse rank of selected genes (rows) in each program (columns). Values indicate the relative importance of these genes to defining a given program. **g**, Pearson correlation of gene expression (rows) and program usage (columns) values in the biopsy samples. Checkpoint receptor genes with high correlation to Program 9 (CD8 T cells) are boxed in black; ligands are boxed in red when involving tumor or vascular programs. **h**, Dot plot of T cell program co-usage within T cell spatial niches in the biopsy samples. Niches are T cell positive bins stratified based on the additional presence of fibroblasts (niche 1), pericytes and endothelial cells (niche 2), or the absence of those stromal components (niche 3; i.e. tumor). Within each T cell niche, program co-usage is calculated for each of 15 programs, setting bins with usage to 1, otherwise 0. The proportion of 1 vs 0 bins are summarized for each niche. **i**, Schematic of the strategy for T cell-proximal gene expression analysis performed in panel (j). Each 24 μm bin positive for a T cell (based on expression of any T cell gene, including (CD3 components, CD4, CD8A, or GCAR) was considered to be within the region of interest, while the remaining bins were considered outside. The radius around each T cell was increased by 1, to encompass up to 3 nearest neighbours. **j**, Proportion of bins within and outside T cell regions in each sample (primary tumor, biopsy cores, xenograft), positive for expression of PD1 (top row) and PDL1 (bottom row). k, Heatmap of Student’s t-test (two-sided) p-values among pairs of regions (n=4 values per region) from panel (j).

To understand why GCAR1 cells were absent from the tumour, we investigated the composition of the TME, aiming to characterize the functional states of the abundant endogenous T cells and their proximal tumor and stromal components. We performed unsupervised discovery of cell types, states, and activities using consensus non-negative matrix factorization (cNMF), with pathway and cell type marker-based annotation of the resulting 15 gene expression programs^27^ (**Methods; Fig. 5c-d; Extended Data Fig. 7a-d**; *Supplementary Tables 5-7* and GitHub repository). This workflow identified five spatially restricted tumor cell activity programs, of which two were differentially abundant between primary and metastatic samples: program 2, characterized by programed cell death (PCD) and insulin signalling predominated in the primary (P2_T_PCD_insulin), while program 5, high in lipid oxidation and Wnt signalling pathways, predominated in the biopsy (P5_T_lipidox_Wnt) (**Fig. 5c**). We identified multiple components of the innate and adaptive immune systems, including myeloid cells, CD4^+^ cells and CD8^+^ cells. In line with IHC results, T cells were highly abundant in the biopsy, were predominantly CD8^+^, and showed signs of exhaustion including PD1 (**Fig. 5c; Extended Data Fig. 7c**). Additionally, multiple stromal cell programs were identified and supported by histology (**Fig. 5c-d, Extended Data Fig. 7c**). Finally, GPNMB expression was highly correlated to spatial usage of tumor programs, indicating that tumor cells were the primary source of antigen (**Fig. 5e**).

To assess possible mechanisms of T cell resistance, we focused our analyses on genes known to modulate immune function, evaluating their relative importance to each program (**Fig. 5f**), and their correlation with program usage (**Fig. 5g**). This revealed tumor cells and the vasculature as distinct sources of immune checkpoint expression. Tumor cells (programs 3, 5) were major sources of the ligands PDL1 (CD274) and PVR (CD155), with the corresponding receptors PD1 and TIGIT being highly expressed on CD8 T cells. Endothelial cells and pericytes/VSMC (programs 6 and 7) produced the TIGIT ligands PVR and NECTIN2, the KLRB1 ligand CLEC2D, and CD80/CD86 (CTLA4 ligands). Stratification of T cell positive 24um bins into niches distinguished T cells in close proximity with vasculature, either with (niche 1) and without (niche 2) fibroblasts, or localized distally from vasculature (niche 3; cellular tumor) (**Fig, 5h; Extended Data Fig. 7f)**. This stratification validated significantly upregulated expression of multiple ligands in the perivascular space (niches1-2; **Extended Data Fig. 7g**; *Supplementary Table 8*). Altogether, our results suggest that there are spatially distinct sources of immune inhibition in the ASPS microenvironment, which could be obstacles to efficacy both at the site of lymphocyte extravasation and within the tumor parenchyma.

Anticipating that GCAR1 and immune checkpoint blockade may be a synergistic combination amenable to future study, we further evaluated expression of the PD1-PDL1 axis in clinical samples and in Patient ASPS-1 xenografts treated with GCAR1. Focusing our analyses on T cell-positive bins, we validated that PD1 expression was specific to T cells, with decreasing signal as more of the surrounding tissue was included, and low expression in regions distal from T cells (**Fig. 5h-i**; *Supplementary Table 9*). PDL1 was similarly upregulated in close proximity to T cells, and low elsewhere in the tissue. Without a pre-GCAR1 treatment biopsy from the same lesion, it was not possible to determine whether engagement of the PD1-PDL1 axis followed GCAR1 treatment or was already established during the patient’s previous round of atezolizumab. However, the significant upregulation of T cell-proximal PD1/PDL1 expression in the xenograft indicated that a single round of GCAR1 treatment is sufficient to trigger this immunomodulatory axis *in vivo* **(Fig. 5j**).

### PDL1 blockade improves GCAR1 therapy in an ASPS xenograft

The PDL1 inhibitor atezolizumab was recently approved for treating unresectable or metastatic ASPS.^28^ That, along with our data showing PD1-PDL1 pathway activation in close proximity to T cells infiltrating a treatment-resistant tumour (**Fig. 5j**), an observation further confirmed by immunohistochemistry (**Fig. 6a**), prompted us to evaluate adjuvant atezolizumab in combination with GCAR1 therapy in preclinical studies. Mice bearing established Patient ASPS-1 xenografts were infused with 1e5 healthy donor-derived GCAR1 cells. This dose chosen because it generated stable disease, PDL1 upregulation, and GPNMB^+^ relapse, similar to what was observed in the GCAR1-refractory biopsy harvested from Patient ASPS-3. Adjuvant atezolizumab initiated four days later generated a complete and durable response in three of four GCAR1-treated mice, providing a significant survival advantage over GCAR1 monotherapy or atezolizumab plus CD19-CAR T therapy (**Fig. 6b, c**). Importantly, atezolizumab potentiated GCAR1 expansion in mice that exhibited a complete response to treatment (**Fig. 6d**).

**Figure 6:**
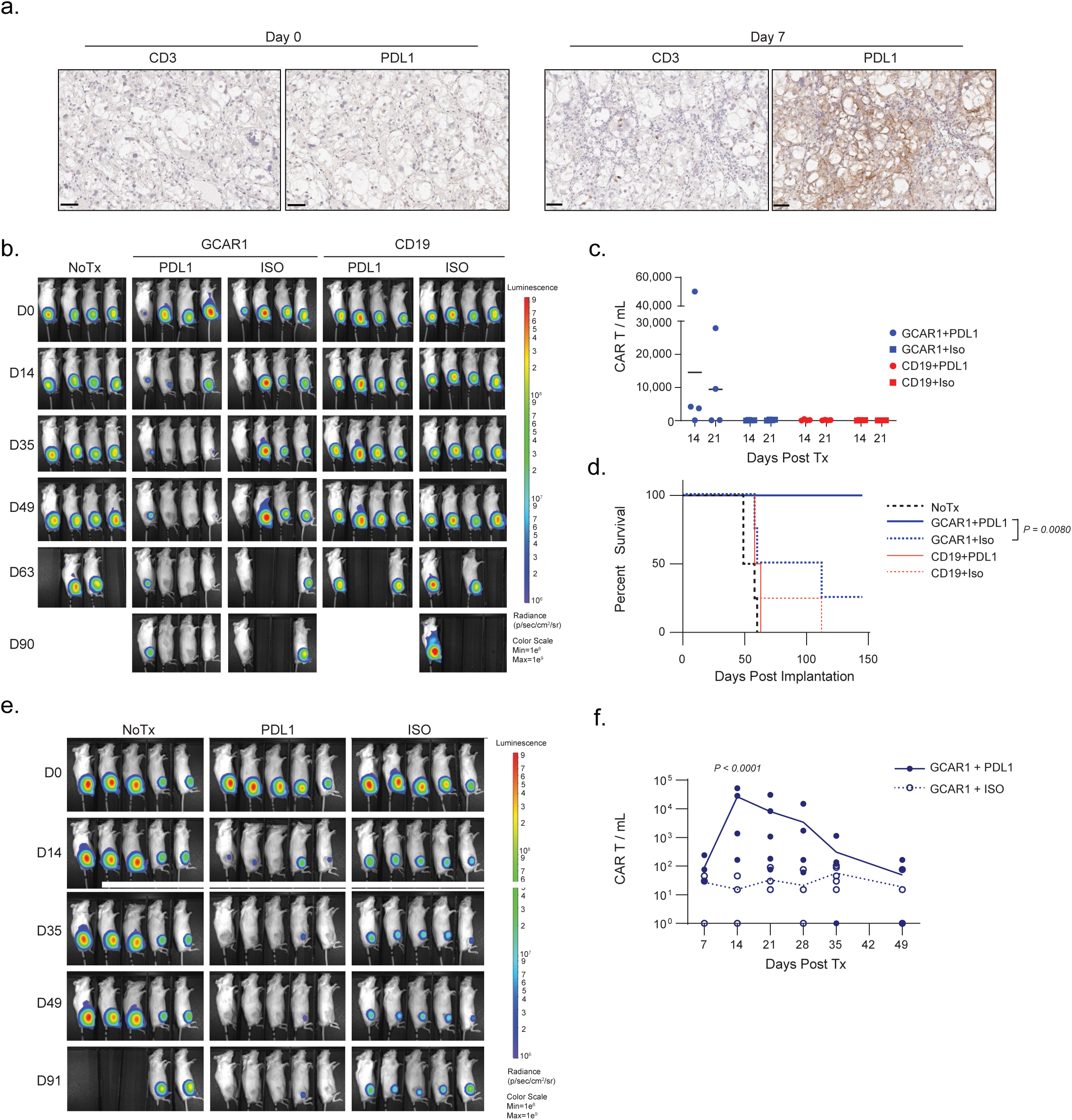
PDL1 blockade improves GCAR1 therapy in an ASPS xenograft. **a**, IHC staining for CD3 and PDL1 expression in tumours resected from ASPS-bearing mice treated with 1e5 GCAR1 manufactured from Patient ASPS-3 T cells. Scale bars = 50μm. **b**, Representative bioluminescent images of ASPS-bearing mice treated with 1e5 healthy donor-derived GCAR1 (n=4) or CD19 (n=4) CAR T cells followed four days later by six doses (2-3 days apart) of atezolizumab or isotype control (ISO). Single experiment. **c**, GCAR1 cells measured by flow cytometry of blood harvested from mice in (b). **d**, Kaplan-Meier survival analysis of mice from (b). (Gehan-Breslow-Wilcoxon test). **e**, Representative bioluminescent images of ASPS-bearing mice treated with 1e5 Patient ASPS-3 GCAR cells (from clinical harvest product) followed four days later by six doses (2-3 days apart) of atezolizumab or isotype control (ISO). Single experiment. **f**, GCAR1 cells measured by flow cytometry of blood harvested from mice in (e). 2way ANOVA with Holm-Šidak’s multiple comparisons test.

To validate this finding and strengthen a justification for retreating Patient ASPS-3 with GCAR1 plus atezolizumab, we conducted a second preclinical study using the clinical trial product previously manufactured for Patient ASPS-3 (*Supplementary Figure 8*). Consistent with the first study, 1e5 GCAR1 cells elicited durable ASPS control when combined with adjuvant atezolizumab (**Fig. 6e**). Marked peripheral expansion was also observed when GCAR1 was combined with atezolizumab (>1500-fold at D14; **Fig. 6f**). Collectively, these data provide a strong rationale for testing GCAR1 therapy in combination with PDL1 inhibition in ASPS patients, via the now standard-of-care atezolizumab.

## DISCUSSION

While CAR T therapies have failed in most patients with solid tumors, recent reports of deep clinical responses in neuroblastoma,^4^ sarcoma,^2^ and brain cancer ^6^ demonstrate that they can be effective, even against difficult-to-treat solid tumours. Here, we provide proof-of-concept for treating a MiT fusion-driven solid malignancy, ASPS, which is incurable once metastatic, with a novel CAR T cell therapy targeting GPNMB.

Although limited to data from a single patient, our study provides a strong justification for testing GCAR1 in more patients with ASPS and other MiT fusion-driven cancers. First, GCAR1 was well tolerated by our patient. Despite robust expansion in peripheral blood, with effector GCAR1 cells comprising >20% of circulating cells at peak expansion, and strong activity against many lung lesions, the patient experienced only mild toxicities, without CRS or ICANS. She did, however, develop an urticarial rash that coincided with peak GCAR1 expansion. Previous studies, and our own observations, have reported GPNMB expression in keratinocytes and melanocytes located in the epidermis.^29^ Indeed, rash was amongst the most commonly reported adverse events in patients treated with a GPNMB-targeting antibody-drug conjugate, glembatumumab vedotin, which was developed using the same scFv as GCAR1.^30–36^ While easily managed in our study, careful monitoring will be required of patients subsequently treated with GCAR1, especially when using escalating doses or in combination with PDL1 blockade, which could dampen peripheral tolerance mechanisms in the skin.

It is also encouraging that GCAR1 expanded and generated measurable anticancer activity in our patient. Indeed, the magnitude, kinetics and phenotype of polyclonal-expanded GCAR1 cells is consistent with the pharmacokinetics of CD19-CAR T cell therapy in patients with leukemia and lymphoma.^37^ That said, her two index lesions were mostly resistant to treatment. This was not unexpected, however, as larger lesions are known to be more refractory to immunotherapy.^38^ It is noteworthy that resistance in the biopsied lesion was not caused by antigen loss, supporting our hypothesis that fusion-driven cell surface proteins with oncogenic function, such as GPNMB, may be less likely to downregulate in response to immune pressure. In contrast, our spatial transcriptomics data suggest that multiple T cell inhibitory pathways, including the PD-1 and TIGIT pathways, may have played a role in mediating treatment resistance. Their expression in multiple distinct spatial niches points toward a multifaceted immunosuppressive response onto GCAR1 cells within the TME. Indeed, immune checkpoint upregulation around tumor vessels suggest that GCAR1 cells may have experienced immunosuppression as early as extravasation, potentially explaining why they were so scarce at 35 days post-treatment.^39^ Fortunately, this might be overcome by infusing more cells or combining with immune checkpoint blockade, possibilities supported by our preclinical data and that we are currently discussing for our patient. It could also be addressed by rewiring GCAR1 cells for resistance to immunosuppressive signaling in the TME, an approach that we are currently exploring in preclinical studies.

In addition to a robust, polyclonal activation of CAR positive T cells, indicating strong GPNMB engagement, we also observed activation of CAR negative T cells after GCAR1 treatment. This has also been reported for other CAR T therapeutics,^2^ and underlies clinical responses to atezolizumab monotherapy in ASPS patients (NCT03141684).^28^ Given that immunogenic neopeptides were predicted for ASPSCR1-TFE3 fusions, it is possible that both GPNMB-reactive CARs and endogenous tumor-reactive TCRs were activated against the ASPS cancer post-treatment. Indeed, this fits our observation of CAR negative CD8^+^ T cells accumulation in the biopsied lesion after GCAR1 therapy. However, as we did not collect a biopsy immediately before GCAR1 treatment, it is possible that these cells were remnants of an unsuccessful response to the patient’s previous course of atezolizumab, in a tumor that had already established a T cell suppressive microenvironment as a counter measure. If so, this suggests that the timing of GCAR1 and atezolizumab within a combination treatment regimen should be carefully considered.

Overall, our data provide a strong justification for studying GCAR1 therapy in more patients with ASPS and other cancers driven by MiT gene fusions. Moreover, fusion-negative cancers harboring other oncogenic drivers of GPNMB, including subtypes of RCC,^17^ triple-negative breast cancer,^40^ melanoma^35^ and osteosarcoma,^30^ may also benefit from GCAR1 therapy. Indeed, a companion manuscript by Savage *et al*. (under review) reports that GCAR1 can target immunosuppressive macrophages in the TME, suggesting that it could have broad utility against many solid tumour types rich in GPNMB-expressing myeloid cells. A Phase I study in patients with ASPS, RCC and TNBC is expected to begin enrolment in 2025.

## ACKNOWLEDGMENTS

ASPS material used for immunohistochemistry originates from multiple European institutions involved in the European Organization for Research on Treatment of Cancer clinical trial EORTC 90101, NCT01524926. We acknowledge the Kelsey Meyer, Amy Abel and staff at the Clinical Research Unit of the Tom Baker Cancer Centre for regulatory and clinical conduct within the single patient study. Figure 2a was designed with www.BioRender.com for which we have an institutional license. This work was supported by the University of Calgary’s Centre for Health Genomics and Informatics, and the University of Calgary’s Applied Spatial Omics Centre.

## AUTHOR CONTRIBUTIONS

FJZ, DJM, MS and ASM conceived and led the study. FJZ, ASM, MS and DJM wrote the manuscript, with input from all authors. ZB developed the regulatory packages and participated in study submission and conduct. JWH, MS, RS, and KH led clinical decision-making and led clinical data interpretation. VL and MJM shared clinical insights. MJM, JAC, and LG2 directed surgical and biobanked biospecimen selection, collection and clinical annotation. DS generated ASPS cell line and xenograft. HL, LS, TL, VN, KE and FJZ conducted in vitro and in vivo preclinical studies. JQ, RH and JB conducted GMP Vector Production. NP, ZB and KP conducted GCAR1 manufacturing and release testing. JBM and PG oversaw genomic and panel sequencing library and data generation. AW and PS provided samples (TMAs, other). JP conducted organoid testing, directed by JAC. AB, CA, JP, BYA generated H&Es, pathology annotations, Visium HD and single cell libraries, directed by JAC, and with contribution by LD. HS, GSG, HAS designed and performed the computational analyses, with contributions from TBV, VTM, KN, AG, MKM, GNYG, directed by ASM, with input from FJZ and DJM.

## DATA AND CODE AVAILABILITY

### Data deposition

Raw and processed single-cell 5’ and V(D)J Enrichment data are deposited at the GEO under accession number GSE283385. Raw and processed data include fastq files and CellRanger output respectively, for 14 samples (7 scRNA-seq and 7 TCR-seq). The raw and processed VisiumHD spatial gene expression data are deposited at the GEO under accession number GSE282057. Raw and processed data include fastq files and SpaceRanger output respectively, for 4 samples. Transcriptome sequencing data and deep sequencing panel data are being deposited in EGA (accession in progress). All other data are included in the main text, the Supplementary Information or are available from the authors, as are unique reagents used in this article. Source data—including the raw numbers for charts and graphs—are available in the Source Data file whenever possible.

### Code availability

Code repository is available on GitHub at https://github.com/MorrissyLab/GCAR1_public, and further includes video files (Extended Data Figure 5b,c) and Supplementary Figure 10.

## FUNDING ACKNOWLEDGEMENTS

DM, ASM, and JC were supported by the Canadian Cancer Society (CCS) [grant number 2020–707161], Financial assistance was provided by generous community donors through the Alberta Children’s Hospital Foundation (ACHF), Alberta Cancer Foundation (ACF) and University of Calgary. DM, MS, FZ and ASM were supported by a Canadian Institutes of Health Research (CIHR) Operating Grant [grant number 400678] and grants from BioCanRx and ACF. ASM was supported by a Canadian Institutes of Health Research (CIHR) Operating Grant [grant number 438802]. ASM holds a Canada Research Chair (CRC) Tier 2 in Precision Oncology. JC was also supported by a Kids Cancer Care Chair in Pediatric Oncology and philanthropic funding from Charbonneau Cancer Institute. FZ and MM are supported by the Kids Cancer Care Foundation. HS was supported by a Marathon of Hope Cancer Centres Network Health Informatics & Data Science Award from the Terry Fox Research Institute [RPGA 3262-09], and a Charbonneau Cancer Institute Postdoctoral Scholarship, awarded by the University of Calgary’s Cumming School of Medicine and the Riddell Centre for Cancer Immunotherapy. GSG was supported by the Alberta Graduate Excellence Scholarship (AGES) and the Katherine Sarah Melinda Mei-Ling Thomas Rare Diseases/Biomedical Engineering Research Scholarship. VTM was supported by a Clark H Smith Brain Tumour Centre Graduate Scholarship. AG was supported by an Alberta Graduate Excellence Scholarship (AGES), a University of Calgary Faculty of Medicine Graduate Council Scholarship, Alberta Innovates Graduate Student Scholarship, and Margaret Rosso Graduate Scholarship in Cancer Research. LM was supported by a Canadian Cancer Society Research Training Award—PhD in partnership with The Terry Fox Research Institute (CCS award #707977 /TFRI grant #1151–06).

## METHODS

### Ethics approval statement

Analyses of human tissue was undertaken according to protocols approved by the Health Research Ethics Board of Alberta (HREBA) Cancer Committee (CC) and in compliance with all relevant ethical regulations. Tumor tissue and patient and healthy donor blood samples were collected with participant consent from the Clark Smith Tumor Biobank at the University of Calgary (HREBA.CC-0762) and for creation of cell lines and use of tissue (HREBA.CC-16-0144). Preclinical studies on human cells and tissue was approved (HREBA.CC-23-0086). Animal experiments and model development were approved by the University of Calgary Health Sciences Animal Care Committee (under protocols AC13-0323, AC21-0016, AC20-0128 and AC24-0135) and complied with all ethical regulations. Apheresis of EHE patient was approved under HREBA.CC-22-0367. The clinical trial protocol was approved by Health Canada (c#276646) and our institutional ethics board (HREBA.CC-23-0151). The patient provided informed consent prior to enrolling on trial. Clinical trial registry number NCT06827886.

### Target discovery pipeline

We downloaded a pre-existing, microarray-based gene expression profile of ASPS^1^ (GSE13433, 14 ASPS samples and two universal RNAs) from GEO. First, we mapped log-transformed and normalized expression values at the probe level to the official gene symbols using the R package *biomaRt* and consolidated the level of gene expression by taking the average across probes mapping to the same gene symbol, which yielded 17,545 genes in total. We then averaged the technical duplicates for each ASPS tumor, *n*=7, before assessing differential expression (DE). We selected 5,725 cell surface proteins (genes) using an integrated database of the surfaceome, where four databases (Cell Surface Protein Atlas,^2^ COMPARTMENTS,^1^ Human Protein Atlas^3^, SURFY^4^) were integrated to prioritize them. Eleven DE genes identified with SurfaceGenie^5^ score >2 were considered as our final candidates. The DE genes were obtained from Welch’s *t*-test between the ASPS tumor and universal RNA groups and false discovery rate (FDR) corrections were applied. Genes were considered DE if FDR < 0.01 and log2-fold-change > 0.5 (i.e., ASPS-specific gene). Lastly, we downloaded two evaluation datasets^6,7^ (GSE32569 and GSE49327) from GEO where there were only ASPS tumor profiles (i.e., pre- or post-treatment, primary or metastasis), and they were processed the same way described above to confirm the level of expression in top candidates. GPNMB expression comparison across different pediatric and AYA tumor was analyzed from whole-transcriptome sequencing analysis of 657 pediatric/AYA extra-cranial solid cancer samples (PMCID: PMC86428100). 37 housekeeping genes were *ACTG1, ACTN4, ACTR1A, ACTR2, ACTR3, AES, AP2M1, BSG, CD59, CSNK2B, EDF1, EEF2, GABARAP, GAPDH, HNRNPA2B1, HSP90AB1, LDHA, LOC100288602, MLF2, MRFAP1, MTCH1, PCBP1, PFDN5, PPIA, PSAP, RAB11B, RAB1B, RAB43, RAB7A, RHOA, RPS27A, SFTPA1, TBP, TUBB, TUBB2A, UBB, YWHAZ*.

### Analyses of GPNMB expression across tissues

We mined two proteomics databases, Human Protein Map^8^ and Wang et al.,^9^ to check GPNMB expression across tissues in non-malignant samples, additionally including CD276 and HER2 as established CAR targets, and 14 housekeeping genes (https://www.genomics-online.com/resources/16/5049/housekeeping-genes/): *ACTB, B2M, GAPDH, GUSB, HMBS, HPRT1, PGK1, PPIA, RPL13A, RPLP0, SDHA, TBP, TFRC,* and *YWHAZ*. Since the range of the mass spectrometry (MS) expression varies and can be extreme, we capped the MS values at the 90th percentile across 17 proteins within a dataset. MS values were then scaled from [minimum, maximum] expression to [0, 1]. Normal tissue microarrays (were obtained from NBP2-78057, NBP2-30170) were obtained from Novus Biologicals and stained as described in immunohistochemistry section.

### Generation of CAR T cells for preclinical studies

Lentiviral plasmids expressing CAR constructs were generated by standard molecular cloning methods. The preclinical GPNMB CAR constructs were assembled from scFv sequences recognizing GPNMB, a MYC epitope tag, a CD8α hinge and transmembrane domain, and 4-1BB and CD3ζ intracellular signaling domains. GPNMB-targeted scFv sequences for G1-G3 were derived from WO2006071441A2 and for G4 from Kuan *et al*.^10^

The preclinical construct was cloned into the pULTRA-EGFP vector (Addgene #24129) downstream of EGFP and separated by a P2A site. The UbC promoter was swapped for a full length EF1α promoter. An identical CD19(FMC63)-targeting construct was also made in this vector. Preclinical lentivirus particles were packaged from LentiX 293T (Takara) cells using packaging plasmid pCMV-dR8.91, envelope plasmid pMD2.G, and CAR construct (5:1:5 ratio). Supernatants containing lentivirus particles were collected 48 after transfection and concentrated by ultracentrifugation. Viral titer in transduction units per milliliter was determined by flow cytometry analysis of transduced Lenti-X 293 or Jurkat cells.

Human PBMCs were isolated from healthy donor or patient blood by Ficol-paque density centrifugation method. CD3 positive T cells were sorted using a CD3 isolation kit (Miltenyi Biotec). Isolated CD3 positive cells were cultured in TexMACs medium (Miltenyi) supplemented with IL7 and IL15 (BioLegend; 10ng/mL) and activated with TransAct CD3/28 beads (Miltenyi). 24hrs after activation, T cells were transduced with CAR-containing lentiviruses at a multiplicity of infection of 5 or 10. Two days later, cells were expanded in ImmunoCult media (STEM CELL) for 8-12 days.

### Cell Lines and *in vitro* CAR Testing

The ASPS PDX-derived cell line was derived from a female with ASPS undergoing surgery for a pulmonary metastases (**Fig. 1e iii)**. Fresh tumor tissue was implanted into the flank of SCID mice and established as a patient-derived xenograft (PDX). Once established, tumour tissue was harvested to create a single cell suspension by gently trituration, from which individual cells were plated in OptiMEM media plus 10% FBS to establish a PDX-derived ASPS cell line. tRCC lines (UOK109, UOK120, UOK124, UOK146 and UOK145) were described previously (REF) and the S-TFE3 line was provided by the RIKEN BRC through the National BioResource Project of the MEXT (Japan), FU-UR-1 was a kind gift from Dr. Kevin Jones, and 786-O, A498 and ACHN from ATCC.

The ASPS-PDX-derived cell line and tRCC lines were transduced with a lentivirus expressing mCherry and luciferase (pLV430G) and sorted for homogenous expression. Stably-expressing cells were subsequently plated in a 96-well plate and treated with indicated ratios of CAR T cells for 24 hours. Cytotoxic activity was quantified by adding luciferin (GoldBio) to a final concentration of 150µg/mL and measured on a SpectraMax i3. IL2 and IFNγ were measured by ELISA kits (BioLegend) and read on the SpectraMax i3.

Explant cultures were created from freshly resected ASPS-2 tumor, cut into ∼1mm^2^ pieces using a scalpel to generate tissue explants. The explants were cultured in organoid medium composed of 1 to 1 mixture of DMEM/F12 (Gibco, cat. no. 11320033) and Neurobasal medium (Gibco, 21103049) supplemented with 1xNEAA (Gibco, cat.no. 11140076), 1% Anti-Anti (Gibco, cat. no. 15240112), 1% N2 supplement (Gibco, cat. no. 17502048), 1% B-27 supplement, minus vitamin A (Gibco, cat. no.12587010), 1x 2-mercaptoethanol (Gibco, cat.no. 21985-023), and human insulin solution (Sigma-Aldrich, cat.no. I9278). The explants were plated in a suspension 6-well plate with 3 ml of organoid medium at the density of ∼20 explants/well and cultured for 7 days on an orbital shaker at 120rpm in a standard cell culture incubator. 50% of organoid media was refreshed daily.

For the co-culture experiment, single explants were transferred to individual wells of a U-bottom 96-well plate containing 150ul of organoid medium with a wide-bore P1000 pipette tip. 1e4 GFP^+^ GCAR1 or GFP^+^ CD19 CAR-T cells derived from ASPS Patient 2 PBMCs were suspended in 150ul ImmunoCult-XF T cell Expansion Medium (Stemcell, cat. no. 10981) and added to the explants for 48 hr. CAR-T cell expansion was visualized with a fluorescent microscope (Echo Revolve; Z-stacks through explant) and the supernatant was collected for the cytokine quantification. GFP mean fluorescence intensity was calculated using Image J v1.54f.

### Animal Models

Six- to ten-week-old female NOD.Cg-*Prkdc^scid^H2-K1^tm1Bpe^H2-Ab1^em1Mvw^H2-D1^tm1Bpe^Il2rg^tm1Wjl^*/SzJ mice (Jackson laboratory; strain #025216) were used in this study. For modelling primary disease, 1e6 ASPS^mC/FLUC^ were injected into the rear leg (biceps femoris). Intracranial metastasis was modelled by steriotactically implanting 1e6 ASPS^mC/FLUC^ into the right striatum. 35-42 days following implantation, mice were treated with the indicated number of CAR T cells intravenously and tumor burden was evaluated by bioluminescence imaging using the Xenogen system and processed with LivingImage Software v4.8.2. For RCC models, 1e6 cells in a 1:1 matrigel:PBS were injected subcutaneously in NOD.Cg-*Prkdc^scid^H2-K1^tm1Bpe^H 2-Ab1^em1Mvw^H2-D1^tm1Bpe^Il2rg^tm1Wjl^*/SzJ (Jackson laboratory; strain #025216). Tumor length (L) and width (W) were measured by calipers, and tumor volume estimated with the following calculation:

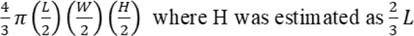

Experimental end-point for survival experiments was reached when tumours grew to ≥15 mm in any direction. CAR T cells were tracked in blood drawn weekly from saphenous vein into EDTA tubes. Animals were monitored daily for general well-being until they reached humane or experimental endpoint.

### PD-L1 Treatment in animals

Animals were treated with an atezolizumab biosimilar (BioXcell, #SIM0009) or Isotype control (BioXcell, #CP171). Mice treated beginning 4 days following administration of GCAR1, and then every 2-3 days for a total of 6 doses. 200ug of antibody was delivered intraperitoneally.

### Flow Cytometry

Single cells derived from cell culture were resuspended in PBS + 2% FBS + 0.25mM EDTA (flow buffer). Alternatively, blood derived from saphenous bleeds was processed in ACK and resuspended in flow buffer. For GPNMB staining of ASPS PDX-derived cell line, anti-GPNMB (R&D MAB25501) with anti-mouse AF647 (BioLegend) was used. The antibodies used for staining T cells are described in the table below. Samples were run on an ATTUNE flow cytometer and data analyzed with Kaluza software. Flow cytometry on samples from the clinical evaluation of GCAR1 was performed on a Cytek Aurora, utilizing the 14-colour Immunophenotyping Panel (Cytek, R7-40000), with of other makers (below). Samples were thawed PBMCs run in parallel from all time points. Data analyzed with Kaluza software. All antibodies used can be found in the Reporting Summary.

### Intravital Microscopy

Tumor IVM was performed as previously described.^11^ In brief, intramuscular mCherry-expressing tumors (∼45 days after implantation) were imaged using a Leica SP8 inverted microscope (Leica Microsystems) on days 2, 4, or 7 post i.v. injection of GFP-expressing CARs (1e6 cells, i.v.). Vessels were counterstained by i.v. injection of 3.5µg CD31 AF647 (BioLegend). Data was processed and analyzed using Leica Las X software and Fiji: Image J. After applying binary masks CARs area was measured and normalized to tumor area. CARs trajectories were analyzed using confinement ratio - the ratio of the displacement of a cell to the total distance that the cell has travelled.^12^

### Immunohistochemistry and Immunofluorescence

Formalin-fixed, paraffin embedded blocks were cut to a thickness of 5µm, and slides were deparaffinized with xylene and rehydrated through an ethanol gradient. Heat-induced epitope retrieval was performed using a microwave with slides in 1X antigen retrieval buffer (10mM Citrate buffer pH 6.0 + 0.05% Tween 20) for 20 minutes at 95°C. Endogenous peroxidase activity was blocked by adding 1 drop of Peroxidase-Blocking Solution (Dako, S202386-2) to each slide and incubating for 15 minutes. Nonspecific binding was blocked with 200 μL of protein block (Agilent, X090930-2) with 0.2 % Triton X-100 (Sigma, Oakville) and incubated for 20 minutes. Each section was incubated for 1 hour or overnight with anti-human GPNMB antibody (R&D systems, AF2550 or Cell Signalling E4D7P, 38313, 1:1000), anti-TFE3 (Sigma-Cell Marque, 354R-15), PDL1 (E1L3N, Cell Signalling), CD3 (SP7, abcam ab16669) or MYC (71D10, Cell-Signaling #2278, 1:200). After washing each section with TBS, 1 drop of the appropriate polymer-HRP secondary antibody (Vector Laboratories or DAKO Envision) was applied, incubated for 30 minutes, washed with TBS, and detected using DAB reagent (DAKO, 1 drop DAB in 1 mL DAB substrate solution). Sections were counterstained with Hematoxylin, then dehydrated through an ethanol gradient, and sections were mounted with Entellan (Electron Microscopy Sciences). The stained slides were scanned using Aperio Scanscope® XT (Aperio Inc.), slide scanner at 20× or 40× resolution and images were acquired, using Imagescope v12.2.2.5015 software or QuPath v0.5.0.

For immunofluorescence, ASPS cell lines were cultured in OPTI-MEM+10% FBS for 48h on chambered slides (Falcon™ 354118) and were fixed with 4% PFA and permeabilized using 0.5% Triston X-100. Each section was incubated with goat polyclonal anti-human GPNMB antibody (R&D systems, AF2550, 20ug/mL) and rabbit monoclonal anti-TFE3 (Sigma-Cellmarque, 354R-15, 1:25) ON at 4°C. The secondary antibody Alexa Fluor® 647 Goat anti-mouse IgG (Biolegend, 405322) was added for GPNMB detection and DyLight™ 488 Donkey anti-rabbit IgG (Biolegend, 406404) was added for TFE3 detection. Both were added at 1ug/mL for 30minutes. Sections were stained with one drop of ProLong™ Gold antifade reagent with DAPI (Invitrogen, P36935). The stained slides were digitalized using ECHO Revolve (ECHO, RVL-100-M) at 40X resolution.

### GMP Vector Production

Clinical GCAR1 plasmid was assembled using the identical GPNMB targeting scFv sequence, hinge domain and MYC epitope as described from the preclinical vector, and then cloned into the CD19-41BBζ-CLIC backbone currently in clinical trial (NCT03765177). Clinical grade GCAR1 transfer plasmid and the accessory plasmids (GagPolRev helper and VSVg envelope) needed for lentivirus manufacturing were produced within the BC Cancer plasmid manufacturing platform, generated under strict, verifiable QA practices as allowed under GUI-0036 (Health Canada Guidance Document Annex 13 to the Current Edition of the GMP Guidelines for Clinical Trials). Each plasmid was sequence verified and individually manufactured aseptically, using a modified alkaline lysis procedure followed by anion-exchange column purification. Manufacturing runs were documented and tracked in Batch Production Records (BPRs). Plasmid release tests included DNA yield (Qubit 2.0), DNA purity (Nanodrop8000 A260/A280 ratio), plasmid identity (fingerprinting by restriction digestion) and Endotoxin (Endosafe PTS). Lentiviral vector was subsequently manufactured at the Biotherapeutics Manufacturing Center (BMC) at the Ottawa Hospital Research Institute. A full-scale GMP run, using a released master cell bank of adherent 293T cells, and consisting of 36 HYPERstacks for a total of 24 L starting material, was conducted in the BMC’s Grade B, CL2 compliant GMP cleanroom. Product-specific release testing was performed at the BMC (identity, potency, and endotoxin) or outsourced to Charles River Laboratories (replication-competent lentivirus, mycoplasma, sterility, appearance, pH).

### GCAR1 Manufacturing

GCAR1 was manufactured from fresh apheresis product collected by leukapheresis using the cMNC procedure on the Terumo Spectra Optia at the Tom Baker Cancer Centre and stored at 4°C overnight prior to manufacturing. GC1 was manufactured using the T cell Transduction (TCT) Standard Protocol on the Miltenyi CliniMACS Prodigy, which starts with an automated CD4/CD8 enrichment by immunomagnetic positive selection and an automatic red blood cell reduction step. From the enriched fraction, 1E8 CD4/CD8^+^ cells were seeded for cultivation and activated with TransACT (anti-CD3/CD28), in Miltenyi TexMACS GMP media, supplemented with 3% GemCell Human AB serum, and reconstituted IL-7 (1,350 IU/mL) and IL-15 (450 IU/mL), per the manufacturer’s directions. Twenty-one hours after activation, cells were photographed for evidence of activation and transduced with 4 vials (2 mL) of GMP lentivirus thawed in a water bath at 38.1°C. Cells were cultured for 11 days at 5% CO_2_ and 37°C, and harvested on Day 12 in 100 mL of final formulation solution containing 25% Alburex (human serum albumin (HSA)) in Plasmalyte-A (PLA). Doses and retains were formulated at 1:1 ratio of drug substance (Harvest Material+HSA) to freeze media (20% DMSO, 5% HSA, PLA) in CryoMACS Freezing Bags (doses) and vials (retains). The cell therapy product (CTP) was placed in a chamber pre-cooled to 0°C and frozen using a standard profile of 1°C drop per min for 60 min followed by 5°C drop per min for 15 min. Upon completion of the controlled rate freezer run, CTP was transferred to LN2 vapour phase freezer for storage. Retained product samples were thawed and release tested for: appearance, pH, total nucleated cell count, pre- and post-thaw viability (7AAD^-^ flow), purity (CD3^+^ flow), immunophenotype (CD4:CD8 by flow), identity (CAR^+^ flow), vector copy number (gag qPCR), potency (cytotoxicity), sterility, endotoxin, and mycoplasma. Testing demonstrated the product was free from microbial contamination and met all predetermined release specifications. Twenty days post cryopreservation, the product was released, thawed at the bedside and administered to the patient.

### ddPCR

Detection and quantification of the Psi sequence integrated into cells following lentiviral transduction were performed using the Bio-Rad QX600 Droplet Digital PCR (ddPCR) system coupled with the AutoDG Droplet Generator (Bio-Rad, California), following the manufacturer’s instructions. Pre-validated, custom-designed primers and probe sets specific for the HIV Psi sequence (FAM-labeled) and the RPP30 reference gene (HEX-labeled) (Catalog Nos. dEXD14812826 and dHsaMDS117591774, Bio-Rad) were used. Genomic DNA was extracted from 5e5 PBMCs using the DNeasy Blood & Tissue Kit (Catalog No. 69504, Qiagen, Netherlands), as per manufacturer’s instructions.

Singleplex ddPCR reactions were prepared in a total volume of 22 µL, consisting of 30 ng of genomic DNA, 1X ddPCR Supermix for Probes (no dUTP; Bio-Rad), 1 µL (5 units) of BamHI-HF restriction enzyme diluted in 1X rCutsmart Buffer (Catalog Nos. R3136 and B6004S, New England Biolabs, Massachusetts), and 250 nM of the corresponding primer and probe mix. Droplet generation was performed using the AutoDG system with microfluidic cartridges and droplet generation oil for probe-based assays (Catalog No. 1864110, Bio-Rad), in accordance with the manufacturer’s instructions.

Thermal cycling was conducted in a PTC Tempo Deepwell Thermal Cycler (Bio-Rad), under the following conditions: initial enzyme activation at 95°C for 10 minutes, followed by 40 cycles of denaturation at 94°C for 30 seconds and combined annealing/extension at 55°C for 1 minute. The final steps included enzyme deactivation at 98°C for 10 minutes and a hold at 4°C for at least 30 minutes. The heated lid was maintained at 105°C, and a maximum ramp rate of 2°C per second was used throughout the cycling protocol.

Following thermal cycling, fluorescence intensity from a minimum of 10,000 droplets per sample was measured using the QX600 Droplet Reader (Bio-Rad). Positive droplets were distinguished using QX Manager Standard Edition software (Bio-Rad), applying manual amplitude thresholds of 1,000 units for FAM and 2,000 units for HEX on 1D amplitude plots to accurately quantify target and reference sequences.

### Volumetric Rendering of CT Images

CT images were taken at Foothills Medial Center. Standard chest thins (0.6-0.625mm) were stacked together in Image J v1.54f and 3D rendered using the Volume Viewer App v2.0. (Mode: Volume, Interpolation: Tricubic Smooth). Images were landmarked between timepoints using vasculature patterns, and xy and xz plains standardized between images. Images of volumertric rendering where imported into Adobe Photoshop 2025, and obvious tumour nodules selected and false-coloured red.

### ASPSCR1-TFE3 targeted breakpoint sequencing (Patient ASPS-3)

Library construction was performed using the KAPA HyperPlus Kit (Roche) with 100ng of genomic DNA. Libraries were quantified and sized by qPCR and a Bioanalyzer 2100 instrument. Libraries were then pooled and hybrid-capture enrichment was carried out using the xGen Hybridization and Wash v2 Kit (IDT) and a custom ASPSCR1-TFE3 panel designed to detect fusion breakpoints. Targeted sequencing (13M reads) was performed on a NextSeq 500 instrument (Illumina) using 76 paired-end reads with 150 cycle mid output kit. Fastq files were merged, and reads aligned to GRCh38 using BWA-MEM with the -M parameter to mark secondary alignments. Reads in the genomic regions chr17:81,977,481-82,017,730 (ASPSCR1) and chrX:49,028,647-49,043,446 (TFE3), and with mapping quality>20 were further analysed. The average read depth in target regions was 59,903. The fusion breakpoint was identified with basepair resolution based on split reads and presence of discordant read pairs. Breakpoints were manually validated in IGV (Integrative Genomics Viewer). SNPs in the flanking regions of the fusion breakpoints were annotated in order to inform subsequent ddPCR primer design.

### Transcriptome sequencing library generation, sequencing, and analysis

RNA library preparations and sequencing were performed by the University of Calgary’s Centre for Health Genomics and Informatics. Input total RNA samples were assessed via TapeStation RNA assay and quantified via fluorescent quantification specific to RNA. Library preparations were performed using Illumina Stranded Total RNA prep with Ribo-Zero plus as per the manufacturer’s protocol. Final libraries were assessed via TapeStation D1000 assay, fluorescent quantification specific to dsDNA and Kapa’s Illumina library quantification qPCR assay. Sequencing was performed on a NovaSeq 6000 S4 300 cycle v1.4 run with paired end 150bp reads, at a target depth of 100M read pairs per sample.

### Single cell and VDJ library preparation, sequencing, and analysis (SPS-Q data)

#### i. Library preparation

##### Cell Preparation

Patient’s PBMCs samples were thawed and washed with warm DMEM + 10% FBS followed by automated cell count. Cells were resuspended in 1X PBS + 1% BSA on a range of 100-2,000 cells/μl. The number of cells loaded on the 10X Genomics Single Cell Chromium Controller was adjusted according to differences in sample concentrations.

##### cDNA amplification and library construction

After generation of single cell gel beads (GEMs), captured cells were lysed, and barcoded full-length cDNA from poly-adenylated mRNA was produced and amplified. An additional amplification was performed with primers specific to TCR constant regions prior to TCR library construction. GEX and TCR libraries were prepared according to the manufacturer’s protocols (10x Genomics Chromium Next GEM Single Cell V(D)J + 5′ Gene Expression).

#### ii. Single-cell TCR (scTCR) sequencing analysis

##### Quality control and normalization

VDJ data was analyzed using *scRepertoire* (v2.0.7).^13^ Each cell was assigned a TCR clonotype based on both TRA and TRB chains. Each TCR clonotype was uniquely identified using a combination of the VDJC genes and CDR nucleotides. Cells with a single TRB chain were retained for further analysis.

#### iii. Single-cell RNA (scRNA) sequencing and analysis

##### Pre-processing and quality control (QC)

Using *cellranger mkref* (v7.0.1),^14^ we generated a customized reference using the aggregated human (GRCh38/hg38) and GCAR1 construct sequences. *cellranger multi* (v7.0.1) was used to align data.^14^ to the Human-GCAR1 customized reference. VDJ^+^ cells from the subset of the GEX dataset were identified and used in downstream analyses. Processing and quality control of the data was performed using *Seurat* (v5).^15^ GCAR1-expressing cells were determined using *‘subset(SeuratObject, subset=GCAR>0)’*. Cells with >10% mitochondrial counts were excluded from the analysis. The GEX data was normalized (*‘NormalizeData()’*). Variable features were selected (*‘FindVariableFeatures()’*) and scaled (*‘ScaleData()’*). After performing Principal Component Analysis (PCA; *‘RunPCA()’*), cells were clustered using *‘FindNeighbors()’*and *‘FindClusters()’* and resolution 2.4, yielding 47 clusters across all samples. Non-linear dimensionality reduction was performed with Uniform Manifold Approximation and Projection (UMAP). Cell doublets were predicted with the embedded Scrublet function in *Scanpy* (v1.10.1), *‘scanpy.pp.scrublet’*. Sample-specific values for doublet thresholds were defined per sample using *mclust* (v6.1.1).^16^

##### Cell type predictions

Unsupervised cell-type labelling was performed with *SCimilarity* (v0.2.0; “*unconstrained*” model).^17^ The resulting 48 cell type labels were rerun using the “*constrained*” model, providing a *‘min_dist’* value for each cell’s label prediction. This quantifies the minimum distance between each query cell and reference cells in the trained model, with 0 being identical. To annotate clusters with cell type information, we calculated a **cluster label score** that incorporates the ‘*min_dist*’ values per label and the number of cells with each label, per cluster, where x is one of the 48 labels, and xc are all cells with label x in a given cluster:

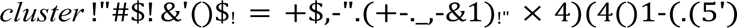

Two of 47 clusters (clusters 29 and 43) had evidence for cell label admixture, and were excluded from downstream analyses.

##### Cluster-level GCAR1 status

This was calculated based on the number of GCAR1^+^ cells, and the proportion of GCAR1 cells. Clusters with >250 or >15% GCAR1^+^ cells were annotated as GCAR1^+^. P-values were calculated to test the significance of the proportion of the GCAR-positive cells in each cluster using the two-proportion z-test (http://www.sthda.com/english/wiki/two-proportions-z-test-in-r).

##### Pathway enrichment analysis

This analysis was performed per cluster using marker genes extracted with ‘performNormalization()’ and ‘FindAllMarkers()’. A subset of each cluster’s marker genes was selected based on the *avg_log2FC >1.0* threshold, and used for multi-query pathway enrichment analysis with the *g:Profiler2* R package (v0.2.3).^18,19^ GO:BP results were used to orthogonally validate the cluster-level cell type annotations.

### GCAR1 probe design

A total of 12 custom spike-in probes for Visium HD were designed against the GCAR1 construct sequence, in accordance with the specifications from 10x Genomics in the: Custom Probe Design for Visium Spatial Gene Expression and Chromium Single Cell Gene Expression Flex Technical Note (CG000621. Rev D; https://cdn.10xgenomics.com/image/upload/v1729114202/supportdocuments/CG000621_CustomProbeDesign_RevD.pdf). Probes demonstrating homology to off-target genes were designed to have at least 5 base pair mismatches in the entire probe sequence. Probe sequences are provided below. The code to generate these probe sequences is available in our Github repository.

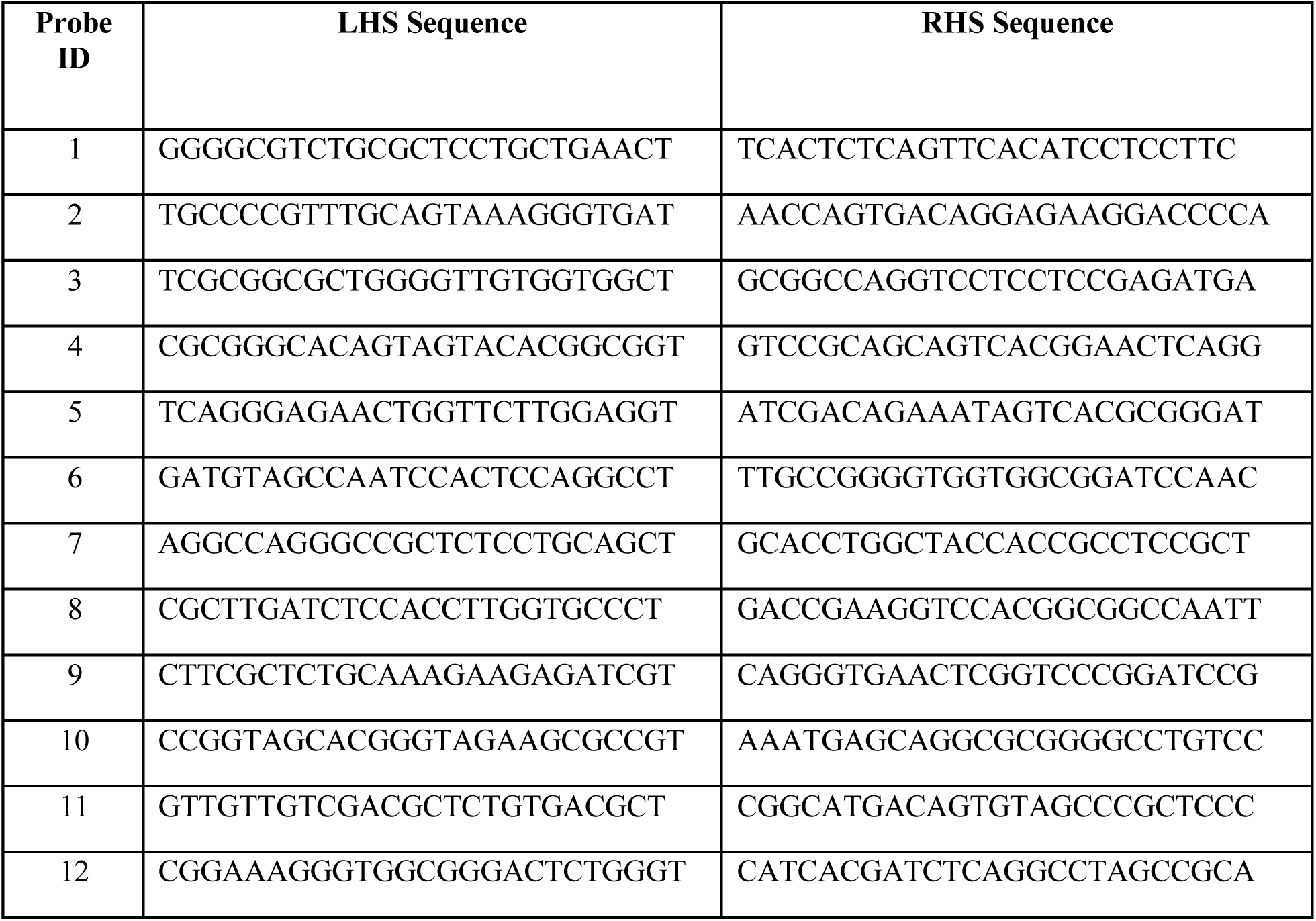

### Visium HD data generation

The Visium HD Spatial Gene Expression Kit (10x Genomics, human probe panel) was used to profile two ASPS biopsy samples, one region of the primary tumor, and a xenograft sample. The GCAR1 probe panel was added on for the biopsy tissues. Tissue samples were sectioned to a thickness of 5 µm and mounted on plain glass slides for deparaffinization, H&E staining, and imaging. A VS110 Slide Scanner microscope (Olympus) at 40x magnification captured tissue morphology before transferring the analytes. To prepare the Visium HD slides, the recommended buffers were thawed, washed, and equilibrated to ensure compatibility with FFPE sections. After imaging, tissue analytes were transferred from plain glass slides to the capture areas of the Visium HD slides (6.5 mm x 6.5 mm) using the CytAssist. The probe extension and library construction steps were performed according to the standard Visium workflow for FFPE outside the instrument. This workflow followed the recommendations in the Visium HD FFPE Tissue Preparation User Guide (CG000684) and in accordance with the Visium HD Spatial Gene Expression Reagent Kits User Guide (CG000685). Sequencing was carried out on an Illumina NextSeq 2000, employing paired-end reads with 43 cycles for Read 1, 10 cycles for i7, 10 cycles for i5, and 50 cycles for Read 2. The NextSeq 2000 generates FASTQ files onboard using Dragen BCL Convert v4.2.7.

### Visium HD data analysis

Using the 10x Genomics SpaceRanger software (v3.0.1 for the primary and xenograft sample; v3.1.1 for the biopsy samples), FASTQ files were aligned against a customized genome reference sequence, comprised of the GRCh38 (GENCODE v44) and the GCAR1 construct sequence. SpaceRanger was used to perform automated image detection and alignment for the primary and xenograft samples, whereas, for the biopsy samples, manual alignment was performed in Loupe Browser (v8.0.0) and exported to be used in SpaceRanger. A custom bin size argument of 24um was passed to SpaceRanger to generate the bin-aggregate data.

We applied unsupervised deconvolution to identify cell types and cell states across all bins. In accordance with previous work, selection of over-dispersed genes was performed per sample using the R package STDeconvolve, with the parameters removeAbove = 1.0, removeBelow = 0.01, and alpha = 0.05.^20^ We utilized consensus Non-negative Matrix Factorization (cNMF) through the mosaicMPI package^20,21^ to identify transcriptional programs across the primary and biopsy samples together and quantify their relative usage within spots. Factorization yielded 15 programs. Program annotation (with cell type, cell states, or pathway activities) was performed using established reference marker genes.^22–24^ Programs that did not demonstrate a match to a cell type or state from the reference marker genes were annotated based on gene set enrichment analysis using gProfiler as previously described, and with results available in our github repository.^18–20^

### Immune checkpoint analysis using Visium HD

#### i. Spatial neighbours

To establish spatial neighborhoods, we first loaded the 24µm bin-size data for each sample (primary, biopsies, and xenograft) into R (v4.3.1) using Seurat (v5.1.0). The samples were then combined into a single Seurat object and filtered through the following steps: (1) merge(), (2) JoinLayers(), and (3) subset(…, subset = nCount_Spatial.024um > 0). Spatial bin coordinates for each sample were extracted from the Seurat object using a custom function, seurat_object_coords(). Neighbors for each bin were identified using the custom program_neighbours() function. To maintain consistency between program deconvolution and gene-based analyses, only bins with a non-infinite normalized usage for any given program were included.

#### ii. GEP neighbourhood niches

Spatial niches were determined by processing the output of the program_neighbours() function. First, bins with usage > 0.05 for Program 9 or Program 10 were identified as bins of interest (BOI). Next, a binarized usage matrix was generated from the cNMF usage output, where usage ≥ 0.05 was set to 1 and all other values to 0. For each BOI and its neighbours at all radii, the mean GEP usage was calculated using the precomputed neighbours table and binarized usage matrix. The resulting data frame was flattened to create a table with BOI as rows and mean program usage at each radius as columns. This data frame was visualized using the ComplexHeatmap R package (v2.16.0), revealing consistent stratification (co-usage) of multiple programs. Based on these, we defined three biopsy niches as follows: bins with presence of fibroblasts at radius 0 (niche 1), presence of pericytes and/or endothelial programs (niche 2), and neither (niche 3). An additional Seurat object was created, containing only the bins displayed in the heatmap. A new column was added to indicate the niche classification for each spot. This refined Seurat object was used to generate the final dot plot via Seurat’s DotPlot() function.

#### iii. T-cell-centric niche analysis

The combined Seurat object from i was subset to only include bins expressing any of the following T-cell genes: CD3D, CD3E, CD3G, CD4, CD8A, or GCAR. The precomputed neighbours table was then filtered to retain only bins expressing these T-cell genes, or BOI. For each radius, neighbouring bins within a given radius of the BOI were classified as either “inside” or “outside.” This classification was stored in the metadata table of the combined Seurat object. Next, the metadata table was merged with the expression levels of genes of interest (PDCD1 and PDL1) from the count matrix. We utilized the following functions from the dplyr R package (v1.1.4) to process the data: (1) group_by(), (2) pivot_longer(), (3) pivot_longer() (repeated for a second operation), (4) group_by(), (5) summarise(), (6) mutate(), (7) mutate() (repeated for another operation), and (8) ungroup(). This pipeline produced a data frame containing the proportion of bins expressing the genes of interest for each condition. The results were visualized using the ggplot2 R package (v3.5.1). Statistical analyses for each condition were conducted using a Student’s t-test, and the nominal p-value exponents were visualized using the ComplexHeatmap package.

#### iv. Between-niche analysis

The radius=0 gene expression status for each biopsy niche was calculated as described previously for T-cell’s. Statistical analyses were conducted on contingency tables generated for each gene, using Fisher’s Exact Test via the fisher.test() function from the R stats package (v4.3.1). Post-hoc comparisons were performed using the pairwise_fisher_test() function from the rstatix R package (v0.7.2). Plotting was done as described above.

### Prediction of ASPSCR1-TFE3 fusion neopeptides and HLA presentation

FASTQ files from patient ASPS-1 and ASPS-4 were aligned with STAR (2.7.11a) and fusions predicted with STAR-Fusion (v1.13.0) using the GRCh38 human reference genome and Gencode v44 annotations from the Trinity Cancer Transcriptome Analysis Toolkit. Candidate fusion junctions were passed to AGFusion (v1.4.1; using the ‘middlestar’ flag) for annotation, and ASPSCR1-TFE3 events were retained for further analysis. HLA alleles for these samples were derived from the WGS data analysis. For samples ASPS-2 and ASPS-3, HLA alleles were typed from patient blood using a commercially available service (CD Genomics). To predict binding to multiple common alleles, a synthetic sample was created that only contained the peptide sequences for the ASPSCR1-TFE3 type I and II fusions, and common HLA alleles. Common MHC class I alleles were shortlisted from the arcasHLA (v0.6.0) HLA frequency database, using the IMGT-HLA database version v3.56.0, and defined as those with an overall population frequency of >0.01. Binding affinities of neopeptides from all samples (including the synthetic sample) were predicted with pVACfuse (v4.2.0) using all available prediction algorithms, and including all peptides with lengths of 8 to 11 amino acids (BigMHC_EL, BigMHC_IM, DeepImmuno, MHCflurry, MHCflurryEL, MHCnuggetsI, MHCnuggetsII, NNalign, NetMHC, NetMHCIIpan, NetMHCIIpanEL, NetMHCcons, NetMHCpan, NetMHCpanEL, PickPocket, SMM, SMMPMBEC, SMMalign). Individual and median IC50 scores (across prediction algorithms producing this statistic) were further analyzed.

### Luminex Analysis

Multiplex cytokine analyses was performed using the Luminex™ 200 system (Luminex, Austin, TX, USA) by Eve Technologies Corp. (Calgary, Alberta). Ninety-six markers were simultaneously measured from blood samples using Eve Technologies’ Human Cytokine 96-Plex Discovery Assay® which consists of two separate kits; the Panel A 48-plex and the Panel B 48-plex (MilliporeSigma, Burlington, Massachusetts, USA). The assay was run according to the manufacturer’s protocol. The Panel A 48-plex consisted of sCD40L, EGF, Eotaxin, FGF-2, FLT-3 Ligand, Fractalkine, G-CSF, GM-CSF, GROα, IFN-α2, IFN-γ, IL-1α, IL-1β, IL-1RA, IL-2, IL-3, IL-4, IL-5, IL-6, IL-7, IL-8, IL-9, IL-10, IL-12(p40), IL-12(p70), IL-13, IL-15, IL-17A, IL-17E/IL-25, IL-17F, IL-18, IL-22, IL-27, IP-10, MCP-1, MCP-3, M-CSF, MDC, MIG/CXCL9, MIP-1α, MIP-1β, PDGF-AA, PDGF-AB/BB, RANTES, TGFα, TNF-α, TNF-β, and VEGF-A. The Panel B 48-plex consisted of 6CKine, APRIL, BAFF, BCA-1, CCL28, CTACK, CXCL16, ENA-78, Eotaxin-2, Eotaxin-3, GCP-2, Granzyme A, Granzyme B, HMGB1, I-309, I-TAC, IFNβ, IFNω, IL-11, IL-16, IL-20, IL-21, IL-23, IL-24, IL-28A, IL-29, IL-31, IL-33, IL-34, IL-35, LIF, Lymphotactin, MCP-2, MCP-4, MIP-1δ, MIP-3α, MIP-3β, MPIF-1, Perforin, sCD137, SCF, SDF-1, sFAS, sFASL, TARC, TPO, TRAIL, and TSLP. Assay sensitivities of these markers range from 0.05 – 100 pg/mL for the 96-plex. Individual analyte sensitivity values are available in the MILLIPLEX® MAP protocol.

### Statistics

Statistical analyses were performed using GraphPad Prism and include paired or unpaired Student’s *t*-test to compare two groups and 2-way ANOVA when analysing multiple groups. Statistical analyses on single cell and spatial transcriptomics datasets were performed in R. Analyses included paired and unpaired Student’s *t*-tests to compare two groups of samples and 2-way ANOVA when analysing multiple groups. A two-proportion z-test was used to test significance of proportions among two groups. Significance on contingency tables was assessed using the Fisher’s Exact test.

## EXTENDED DATA FIGURE LEGENDS

**Extended Data Figure 1:**
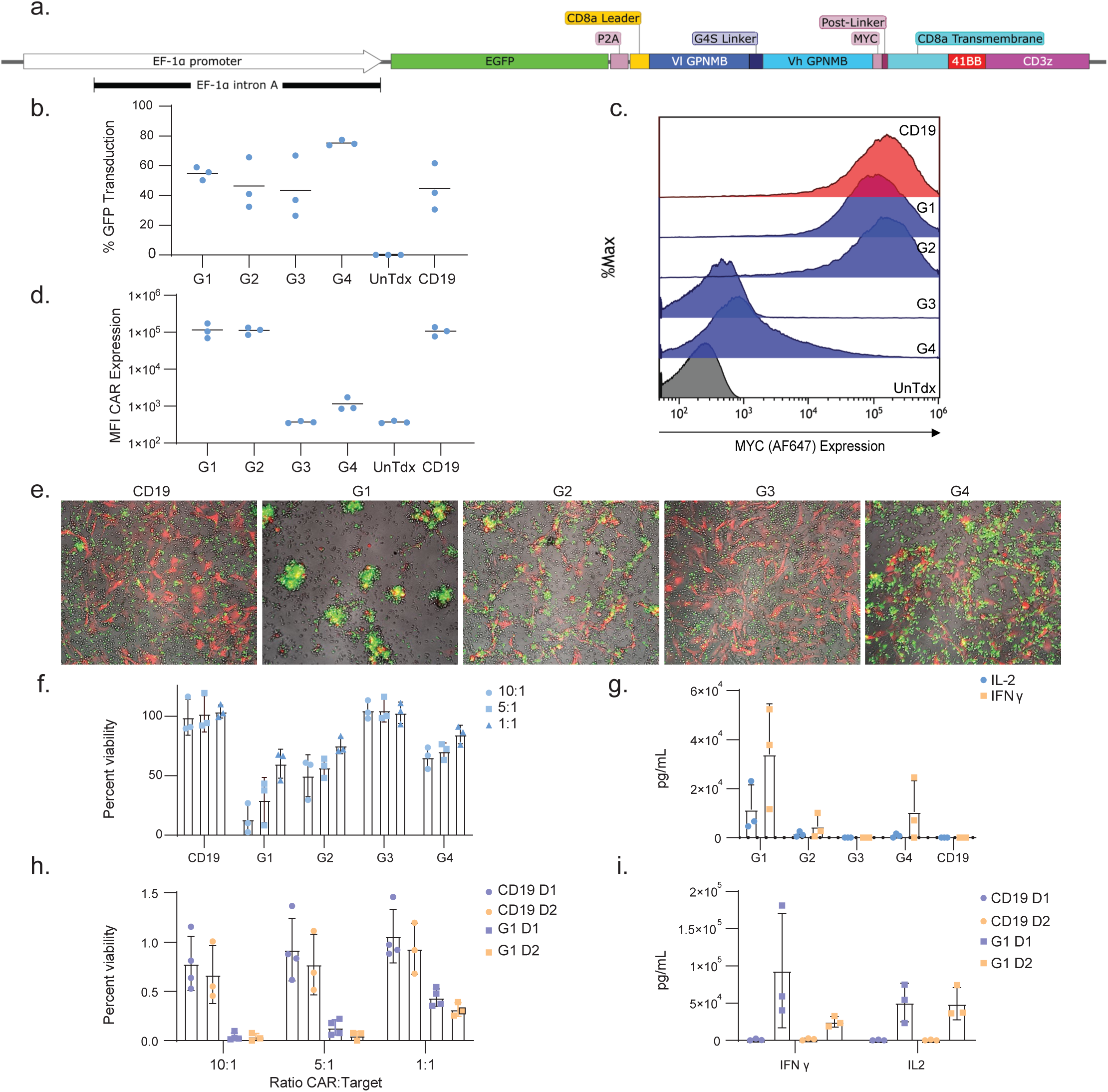
Determining optimal GPNMB Binder for GCAR1. **a**, Components of preclinical CAR backbone from which 4 different GPNMB scFv were tested. Entire construct was cloned into the pUltra lentiviral vector and packaged in a 2nd generation packaging system for preclinical experiments. **b**-**g,** Separate LV packaging and transductions (n=3) were performed on fresh blood draws from the same healthy donor with 10 MOI of virus. Transduction efficiency was measured by looking at the percent positive GFP (c) and CAR expression was assessed looking at median fluorescence intensity (MFI) of MYC in GFP^+^ Cells (d,e). GPNMB CARs were tested for cytotoxic potential on a GPNMB^+^ ASPS PDX-derived cell line (described in *Supplementary Figure 3*). Microscopy images demonstrating an array of cytotoxic potential where the most active GFP^+^ (green) CAR T cells clustered and killed the ASPS cell line (red; e). This cytotoxicity was measured by luminescent assay at listed ratios of CAR T to ASPS target cells compared to a non-targeting CD19 CAR or transduced cells (e) and IFNγ (orange) and IL-2 (blue) secretion of GCAR1 and CD19 CAR T cells at 1:1 ratio in media measured 24 hours after co-culture in vitro on the ASPS PDX-derived cell line (g). **h-i**, Cytotoxicity and cytokine secretion assays were repeated with the lead candidate GPNMB binder (G1) in two other healthy donors (D1 and D2). n=3 of three fresh blood draws and CAR transductions. All bar graphs are means with standard deviation.

**Extended Data Figure 2:**
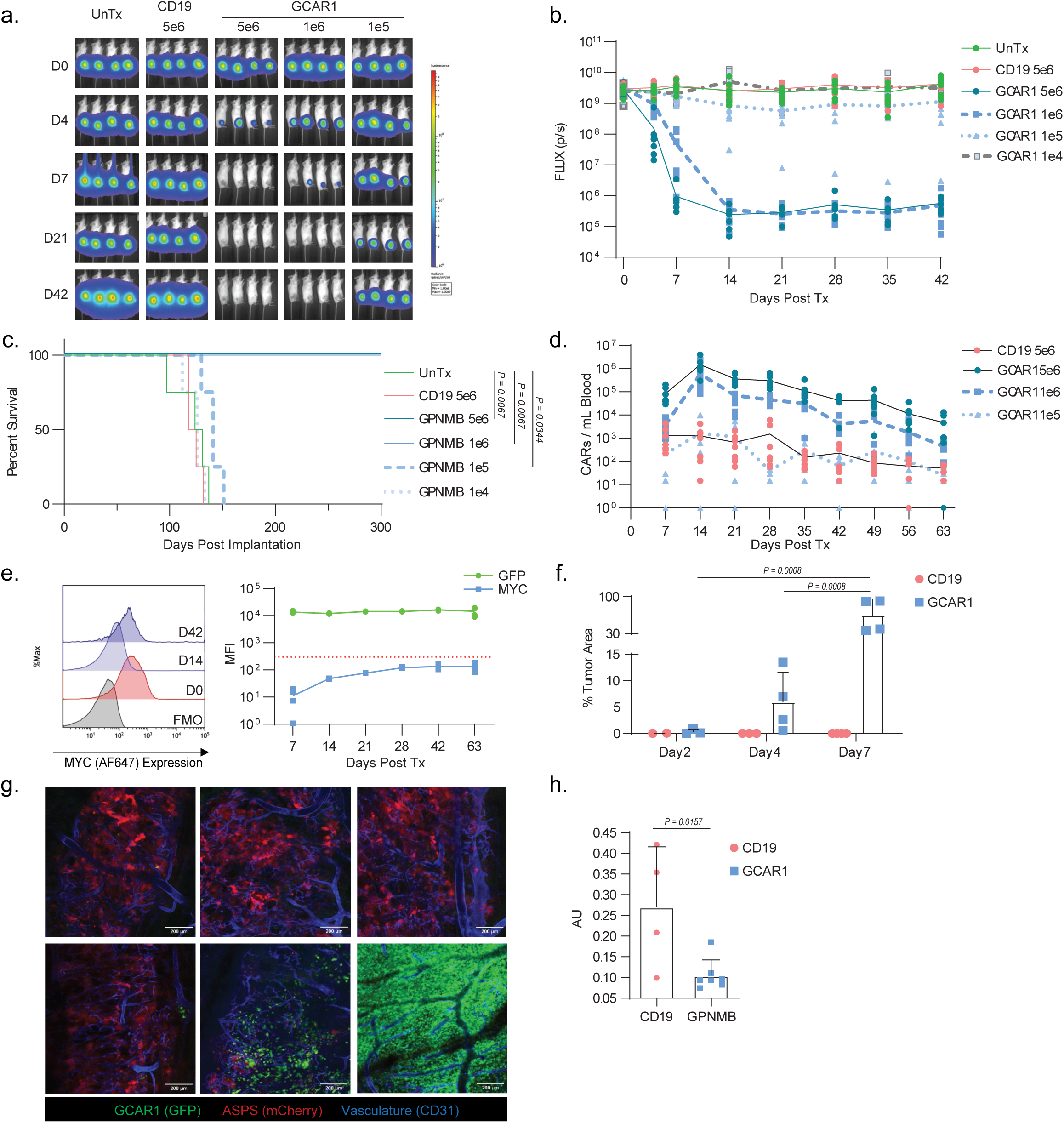
In vivo activity of GCAR1 in mouse model of primary ASPS. NSG-MHC^KO^ animals bearing 42-day intramuscularly implanted ASPS^mc/FLUC^ xenografts (Day 0), and treated with 1e5 (n=8), 1e6 (n=7), or 5e6 (n=7) GPNMB-CAR, 5e6 CD19 CAR (n=7) or untreated (n=8). Results from two independent experiments using two different healthy donor GCAR1. **ab**, Bioluminescent imaging and quantitation of firefly luciferase of ASPS tumours from one healthy donor (n=4). **c,** Kaplan-Meier survival analysis (treatment day 42; Gehan-Breslow-Wilcoxon test). **d,** Measurement of CAR T expansion in the blood with flow cytometry for CD3+GFP+ CARs. 5e6 and 1e6 GCAR1 are significantly different (p<0.001) from CD19 at day 14, and 5e6 GCAR1 is significantly different from CD19 at day 21 (p=0.0141). 2way ANOVA with Holm-Šídák’s multiple comparisons test. Readings of 0 were changed to reading of 1 (10^0^) for the purpose of logarithmic graph. **e,** Merged histogram of all samples (n=4; left) and quantitated mean fluorescent intensity (MFI) flow values (right) of GCAR1 cells in the blood of animals in the 5e6 dose group. D0 represents cell *in vitro* before injection. Blue line represents MYC expression, the green line is EGFP expression, and the dotted red line corresponds to preimplantation MYC expression **f,** Intravital microscopy quantitating GFP^+^ CAR T cells (CD19 red; GCAR1 blue) as a percent tumor area in ASPS tumors 2-, 4-, or 7-days following administration of 1e6 CAR T cells. 2way ANOVA with Holm-Šidak’s multiple comparisons test. Bar graph is mean and standard deviation. **g,** Representative images from intravital microscopy imaged for ASPS xenograft (red), CD31 (vasculature; blue) and CAR (GFP; green). Pictures are stitched images of multiple Z-stacked images (∼100uM depth). i, Analysis of CAR T cell trajectories observed over a span of 20 minutes. Bar graph is mean and standard deviation. Unpaired T test.

**Extended Data Figure 3:**
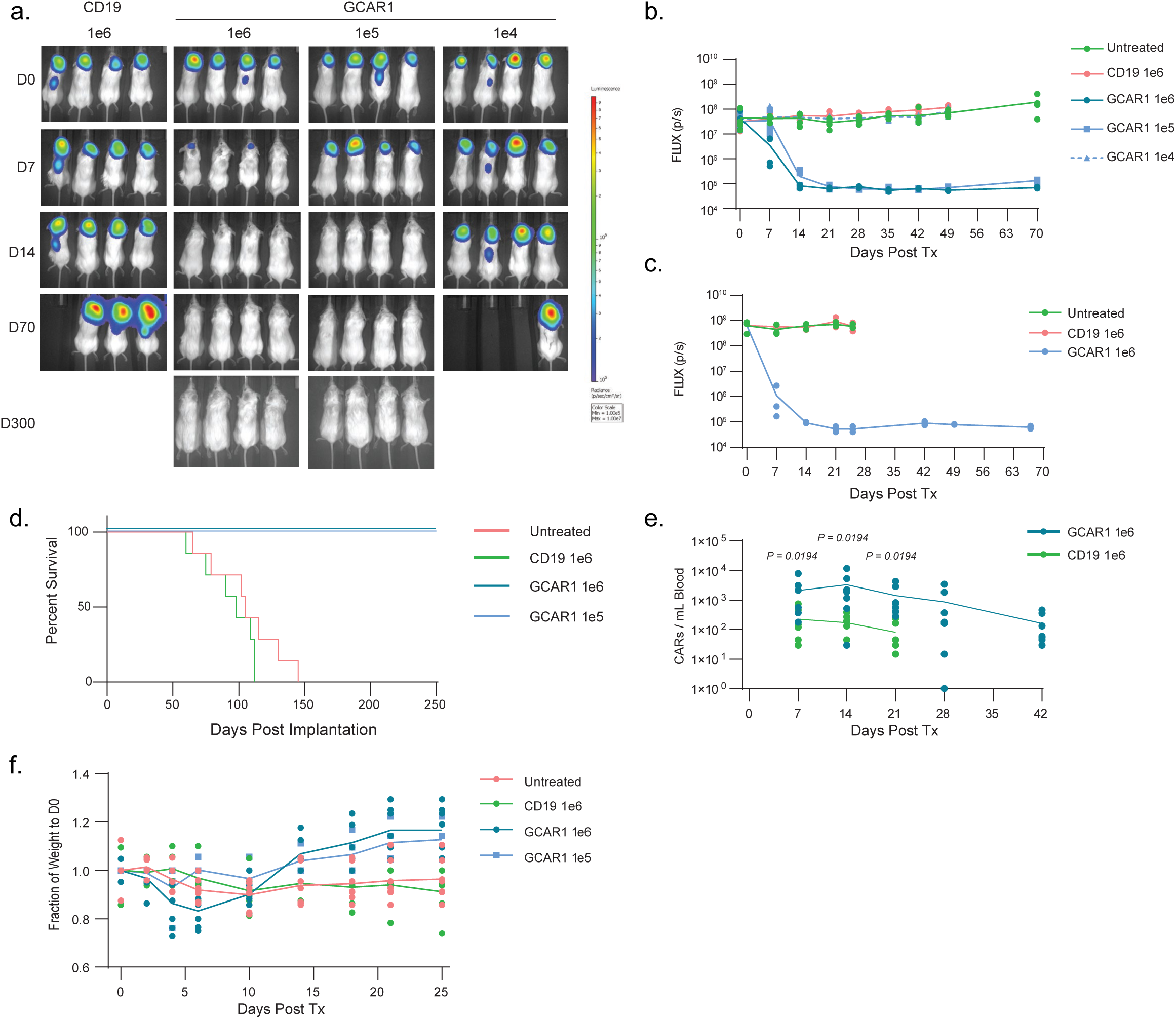
In vivo activity of GCAR1 in mouse model of ASPS CNS Metastasis. **a-b**, Bioluminescent images and quantitation of NSG-MHC^KO^ animals bearing 42-day intracranially implanted ASPS^mc/FLUC^ xenografts (Day 0), and treated with 1e4 (light blue dotted triangles, n=4), 1e5 (light blue solid square, n=4), or 1e6 GCAR1 (dark blue solid circle, n=4), 1e6 CD19 CAR (red solid circle, n=4), or untreated (green solid circle, n=4) and imaged on indicated days. **c,** Repeated experiment different healthy donor with 1e6 GCAR1 (blue solid circle, n=3), 1e6 CD19 (red solid circle, n=3) or untreated (green solid circle, n=3). **d,** Kaplan-Meier survival analysis of both experiments with two healthy donors for untreated (n=7), 1e6 CD19 (n=7) and 1e6 GCAR1 (n=7). 1e5 GCAR1 was only done with one healthy donor (n=4). **e,** Measurement of CAR T Expansion in the blood with flow cytometry for CD3^+^GFP^+^ CARs following weekly blood draws. 2way ANOVA with Holm-Šídák’s multiple comparisons test. Readings of 0 were changed to reading of 1 (10^0^) for the purpose of logarithmic graph. **f,** Weights of animals taken following CAR administration and presented as a percent weight at treatment Day 0. Data is a combined result of the two experiments for untreated (n=7), 1e6 GCAR1 (n=7) and 1e6 CD19 CAR (n=7), and one experiment (n=4) for 1e5 GCAR1 CAR. 1e6 GCAR1 is statistically different than untreated at days 6, 14, 18, 21 and 25 (p<0.05), and 1e5 GCAR1 is statistically different from untreated at day 25 (p<0.05). Two-way ANOVA with Dunnett’s multiple comparisons test.

**Extended Data Figure 4:**
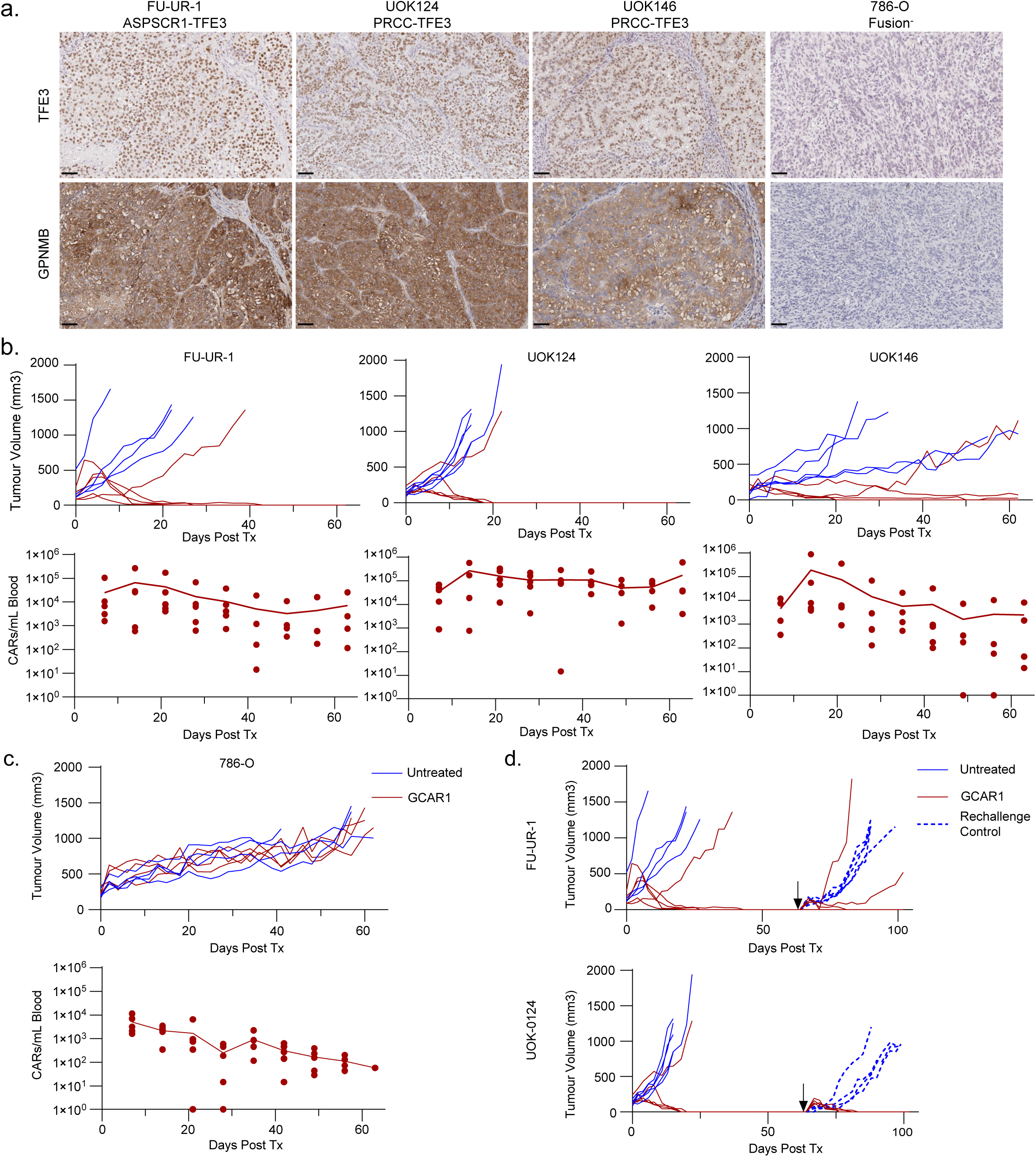
Clinically manufactured GCAR1 is functional in models of translocation-positive renal cell carcinoma. **a**, TFE3 and GPNMB IHC staining of tRCC (FU-UR1, UOK124, UOK146) and RCC (786-O) xenografts subcutaneously grown in NSG-MHC^KO^ mice. Scale bars = 50 um. **b**, Subcutaneous tRCC models were treated with 5e6 clinically manufactured GCAR1 (HD1) cells administered intravenously. Tumor measurements (upper; red line - GCAR1; n=5), blue line - untreated; n=5) and weekly blood draws for measuring circulating GCAR1 cells (lower). Lines are mean of all samples. **c**, Subcutaneous RCC (786-O) model was treated with 5e6 clinically manufactured GCAR1 (HD1) cells administered intravenously. Tumor measurements (upper; red line - GCAR1; n=5, blue line - untreated; n=5) and weekly blood draws for measuring circulating GCAR1 cells (lower). Line is mean of all samples. **d**, Mice that demonstrated complete responses from the FU-UR1 and UOK-124 groups, or naive mice (rechallenge control), were rechallenged with the same tumors and measured for tumor growth.

**Extended Data Figure 5:**
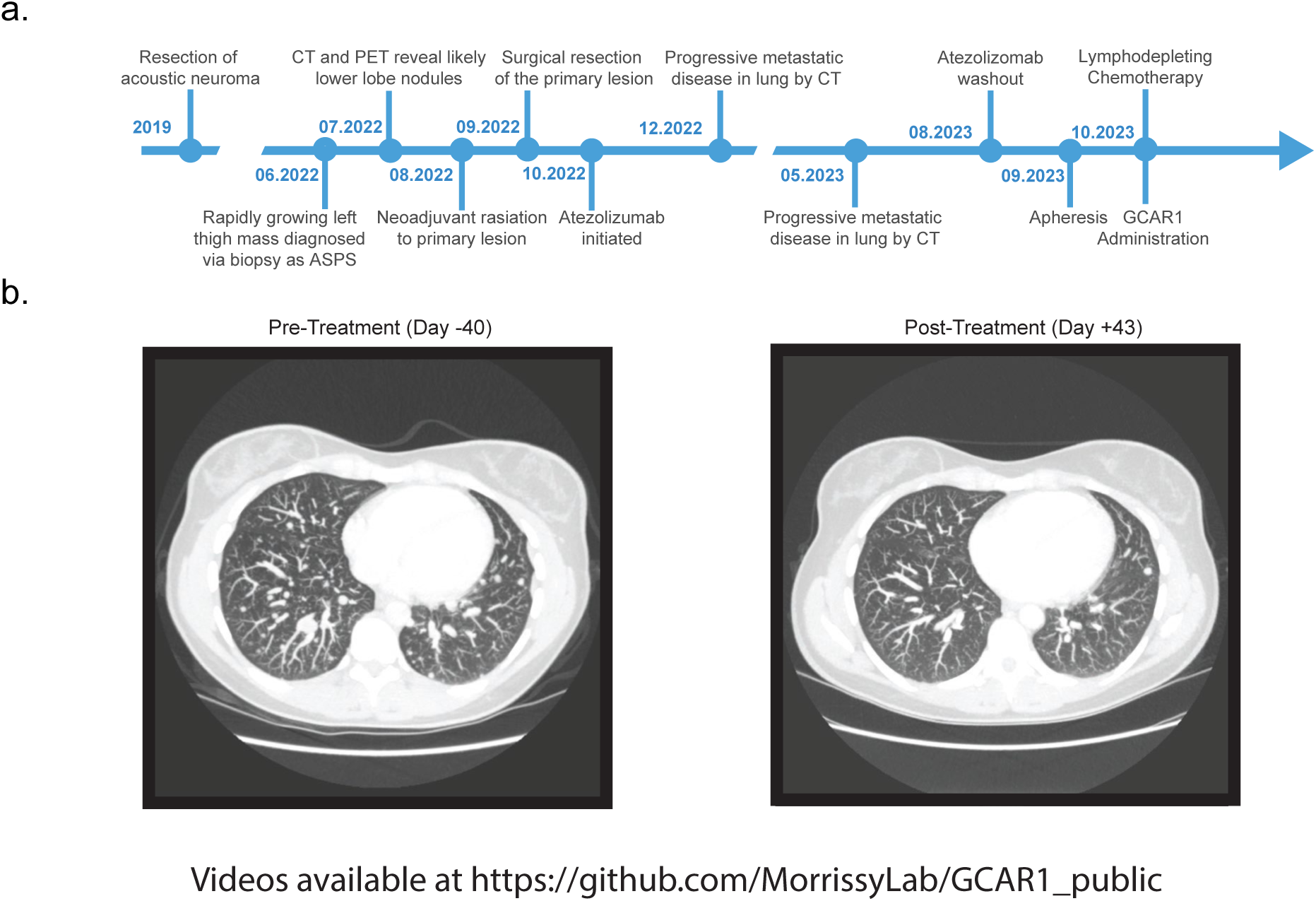
Patient history and treatment course. **a,** The presented with a rapidly growing left thigh mass biopsy-proven to be alveolar soft part sarcoma (ASPS Patient-3). Stage CT and PET scans revealed multiple bilateral lung nodules suspicious for metastatic disease. Following neoadjuvant radiation to the primary site of disease she underwent surgical resection of the thigh mass followed by initiation of atezolizumab. Repeat CT imaging 3 and 9 months later while on atezolizumab confirmed rapidly progressive disease with innumerable new and enlarging pulmonary metastases. Her past medical history was otherwise unremarkable save for a benign acoustic neuroma resected years earlier. Washout The patient received lymphodepleting chemotherapy consisting of fludarabine 40 mg/m2 (Day -4, -3, -2) and cyclophosphamide 600 mg/m2 (Day -3, Day-2), followed by one rest day with a minimum of 48 hours before GCAR1 infusion. A single dose of cryopreserved, autologous GCAR1 was given intravenously at a dose of 1x106 CAR^+^ cells/kg on Day 1 and the patient was subsequently monitored in hospital for any adverse events (AEs). b, Compiled maximum image projection (MIP) images from CT images pre-treatment (day -40) and post-treatment (day +43) in Z-stack video. Note - see attached videos https://github.com/MorrissyLab/GCAR1_public.

**Extended Data Figure 6:**
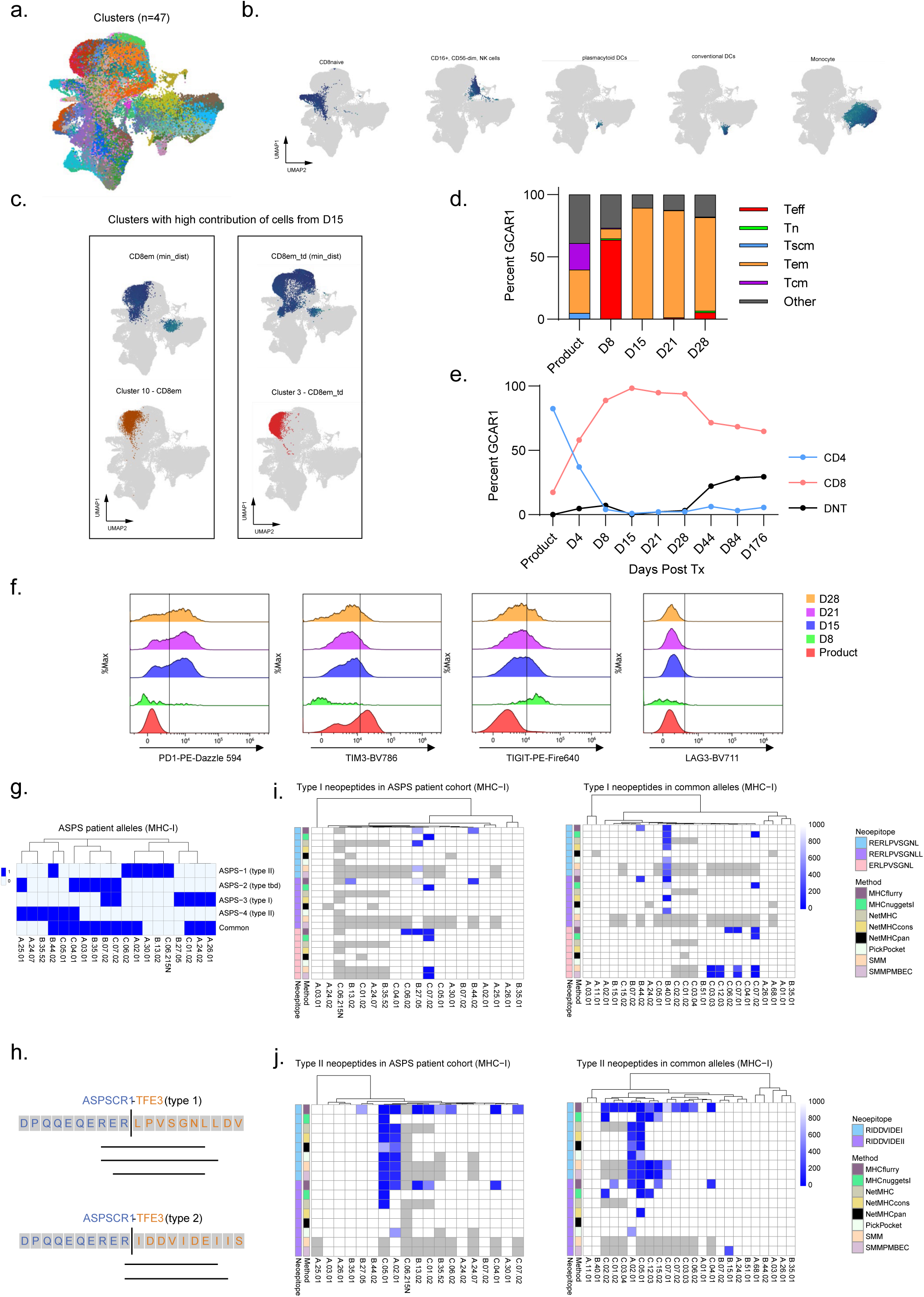
Peripheral expansion of GCAR1 cells, and fusion-derived neoantigens. **a**, UMAP plot of the merged single-cell RNAseq data, annotated with the identity of each of 47 clusters. **b**, UMAP plots showing cell-level annotations predicted by SCimilarity for a subset of cell types. Colour denoted the confidence of each label, with 0 corresponding to maximum confidence (min_dist; see Methods). Scale as in Figure 4f. **c,** UMAP plots showing cell label prediction (SCimilarity) and cluster-level cell class annotation for the most highly prevalent cell classes at D15 (CD8em and CD8em_td). **d,** Flow cytometry analysis of circulating GCAR1 memory phenotypes looking at proportion of naive T cells (Tn; CD45RA^+^CD45RO^-^CCR7^+^CD27^+^CD127^+^CD95^-^), effector T cells (Teff; CD45RA^+^CD45RO^-^CCR7^-^ CD27^+/-^CD127^-^CD95^+^), effector memory cells (Tem; CD45RA^-^CD45RO^+^CCR7^-^CD27^+/-^CD127^+/-^CD95^+^), central memory T cells (Tcm; CD45RA^-^CD45RO^+^CCR7^+^CD27^+^CD127^+^ CD95^+^), stem cell memory T cells (CD45RA^+^CD45RO^-^CCR7^+^CD27^+^CD127^+^CD95^+^) and other (did not fit any of these populations). Cells gated on Live, CD45^+^CD3^+^MYC^+^CD19^-^CD14^-^CD16^-^ cells. **e,** CD4 and CD8 percentages of GCAR1 from preinfusion product to day 28 post-infusion. Cells gated on Live, CD45^+^CD3^+^MYC^+^CD19^-^CD14^-^CD16^-^ cells. **f,** Flow cytometry of expression of T cell inhibitory receptors PD1, TIM3, TIGIT and LAG3 on GCAR1 cells on samples in the ASPS-3 GCAR1 product and circulating GCAR1 cells. Cells gated on Live, CD45^+^CD3^+^MYC^+^CD19^-^CD14^-^CD16^-^ cells. **g,** HLA MHC class-I allele typing results for ASPS patients in the cohort. **h,** Schematic indicating the subset of fusion-spanning neopeptides in the 8-11 amino acid length that were predicted to bind to at least one MHC-I allele in the patient cohort or to common alleles in the general population. These neopeptides had IC50 values < 500 from 3 or more prediction algorithms, and an overall median IC50 value < 1000. **i-j,** Heatmap of binding affinities (IC50) for each peptide in panel (h) (rows) to the set of alleles in 4 ASPS patients (left panels), or common alleles in the general population (>1%; right panels).

**Extended Data Figure 7:**
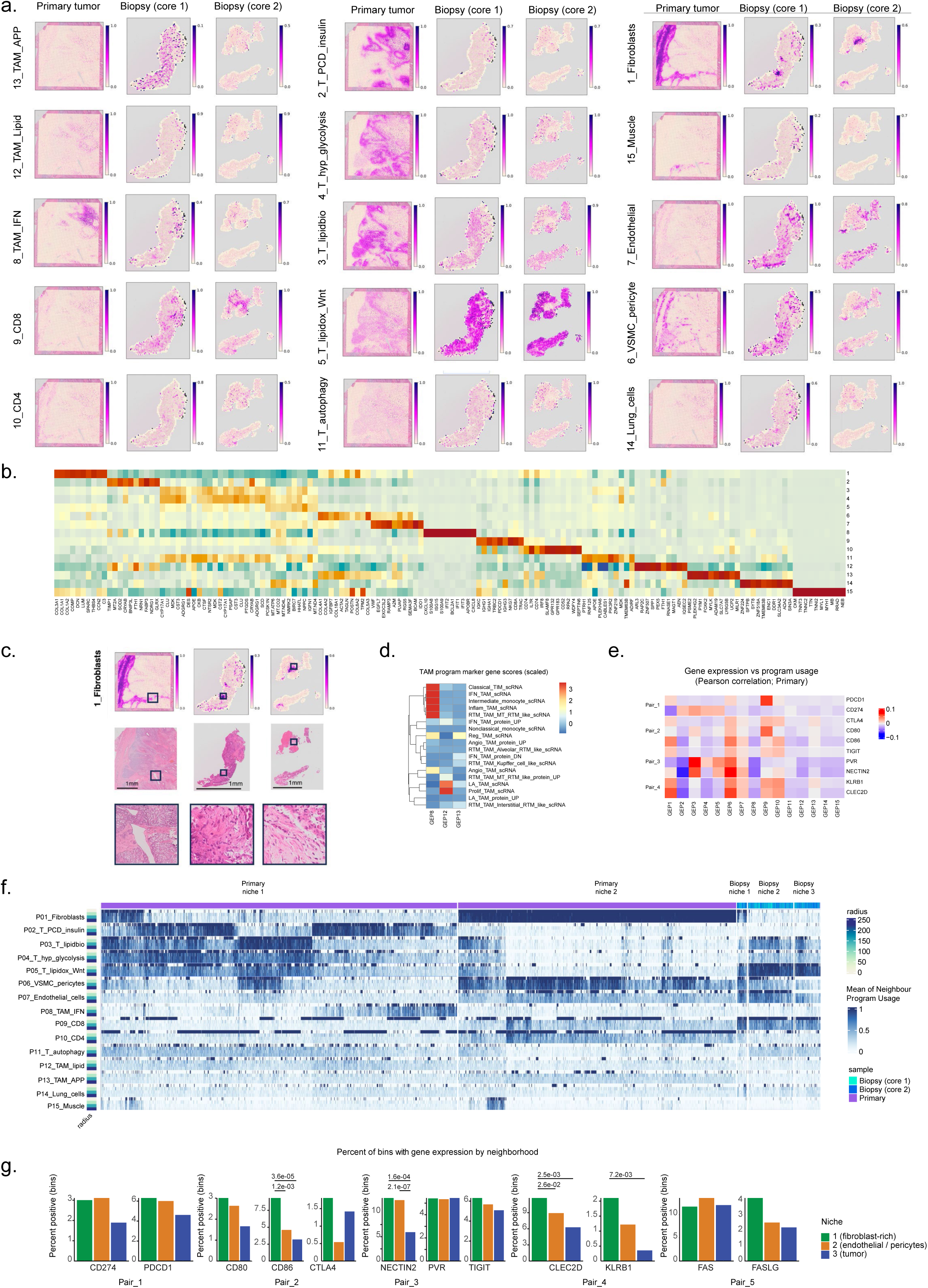
GCAR1 and T cell activity within a GCAR1-resistant lesion. **a,** Spatial plots of program usage for all 15 annotated programs identified across the primary and biopsy samples in Patient 3. **b,** Heatmap of the top 10 most highly scoring (i.e. programdefining) genes (columns) in each of 15 programs (rows). Inverse rank values are center scaled. **c,** Gene expression program 1 (fibroblast) usage and histology for all samples, with histology closeup shown for highlighted regions (black boxes). **d,** Heatmap of scaled marker gene scores corresponding to multiple tumor associated macrophage gene sets (rows) for a subset of TAM programs (columns). **e,** Pearson correlation of gene (rows) expression and program (columns) usage values in the primary samples, to identify correspondence between program usage and gene expression. **f,** Spatial neighbourhood distinction in the biopsy and primary tumor samples (top column). Binarized program co-usage was performed for T cell positive bins (based on program 9 and 10 positivity), and at 3 radii as described in Figure 5i. Hierarchical clustering of the matrix was followed by splitting along two major branches in the primary sample. Within the biopsy, 3 niches were identified as described in Figure 5h, based on co-usage of fibroblast (1) and vascular programs (6, 7). **g,** Barplots of gene-positive bins (percent positive bins on y-axis), for each biopsy T cell niche, including checkpoint receptors and ligands. P-values are calculated using a fisher-exact test (two-sided) on the number of gene-positive versus negative bins in each niche. Sample numbers and p-values are available in *Supplementary Table 8*.

## SUPPLEMENTARY FIGURE LEGENDS

**Supplementary Figure 1:**
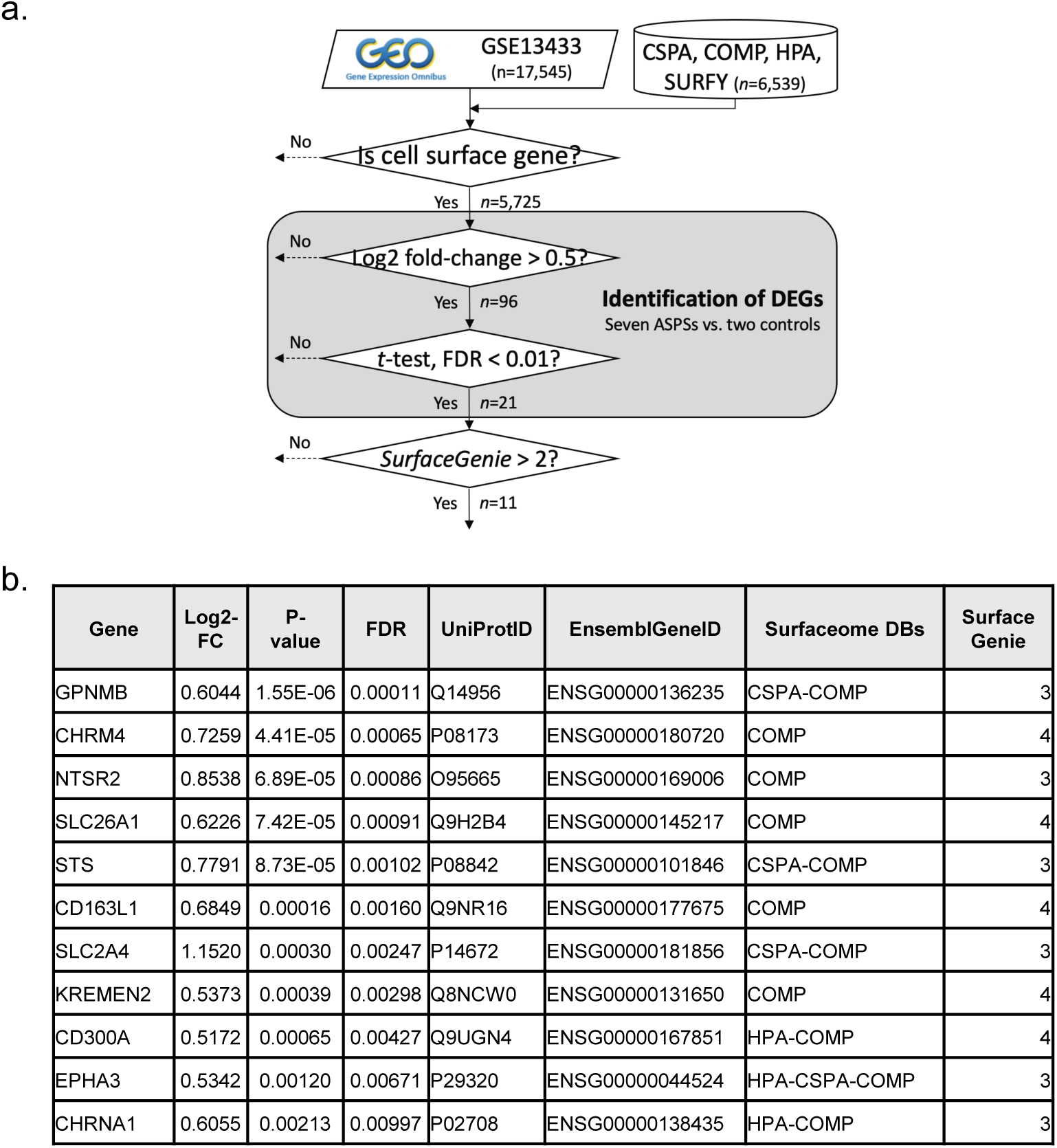
Target discovery ASPS Cell Surfaceome. **a,** Bioinformatics pipeline to identify potentially overexpressed cell-surface proteins in ASPS vs. Normal tissue from microarray data in GSE13433. **b,** Expression of 11 candidate cell-surface ASPS genes derived from the pipeline.

**Supplementary Figure 2:**
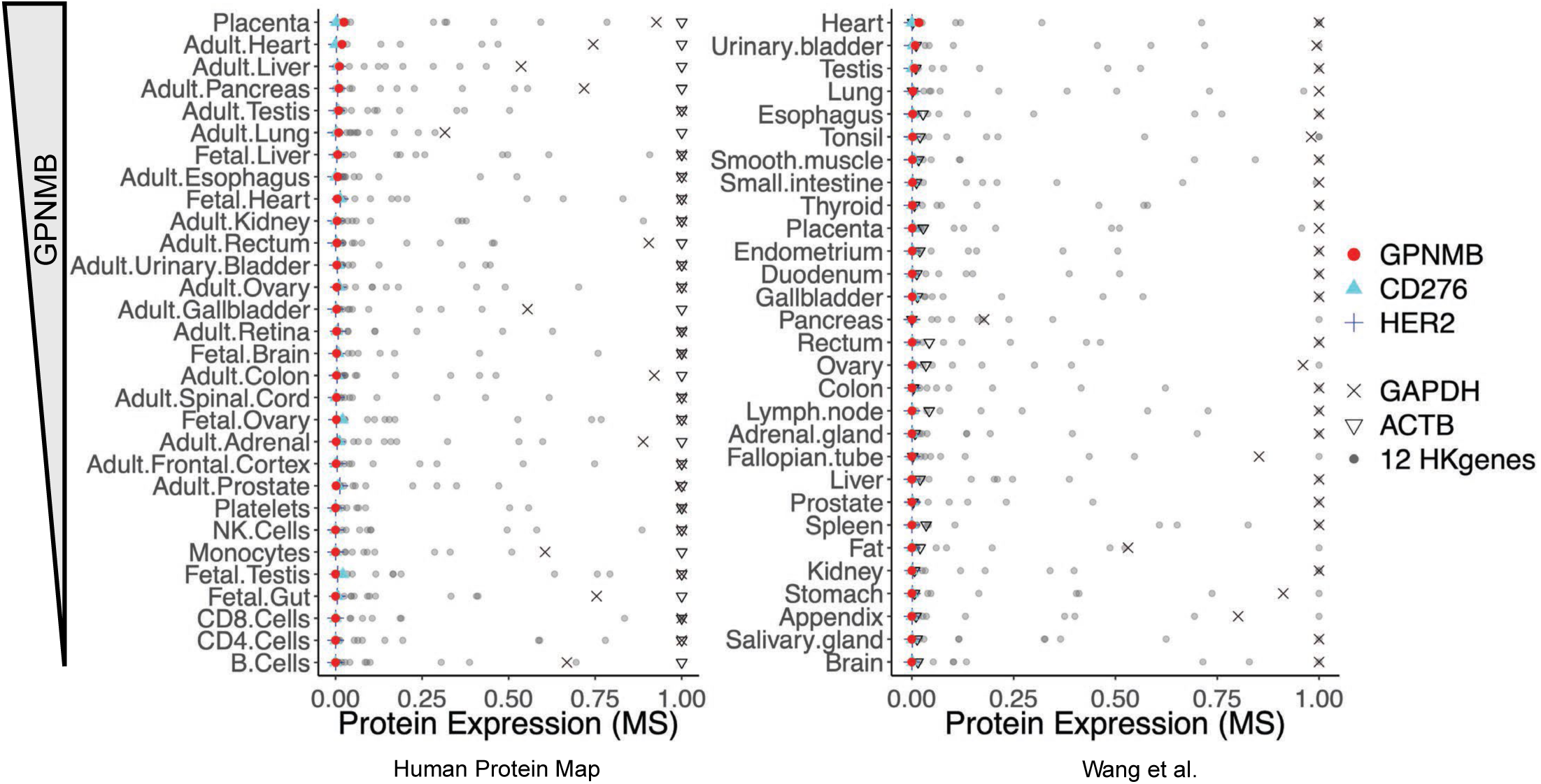
Normal Proteomics DB for GPNMB. Human Protein Map and Wang et al., were used to assess the level of GPNMB (red circle) expression across tissues in non-malignant samples. In addition, we included CD276 (light blue triangle) and HER2 (blue cross) as established CAR targets and 14 important housekeeping genes (https://www.genomics-online.com/resources/16/5049/housekeeping-genes/): ACTB, B2M, GAPDH, GUSB, HMBS, HPRT1, PGK1, PPIA, RPL13A, RPLP0, SDHA, TBP, TFRC, and YWHAZ. Since the range of the mass spectrometry (MS) expression varies and can be extreme, we capped the MS values at the 90th percentile across 17 proteins within dataset. Then, the MS values were scaled from [minimum, maximum] expression to [0, 1].

**Supplementary Figure 3:**
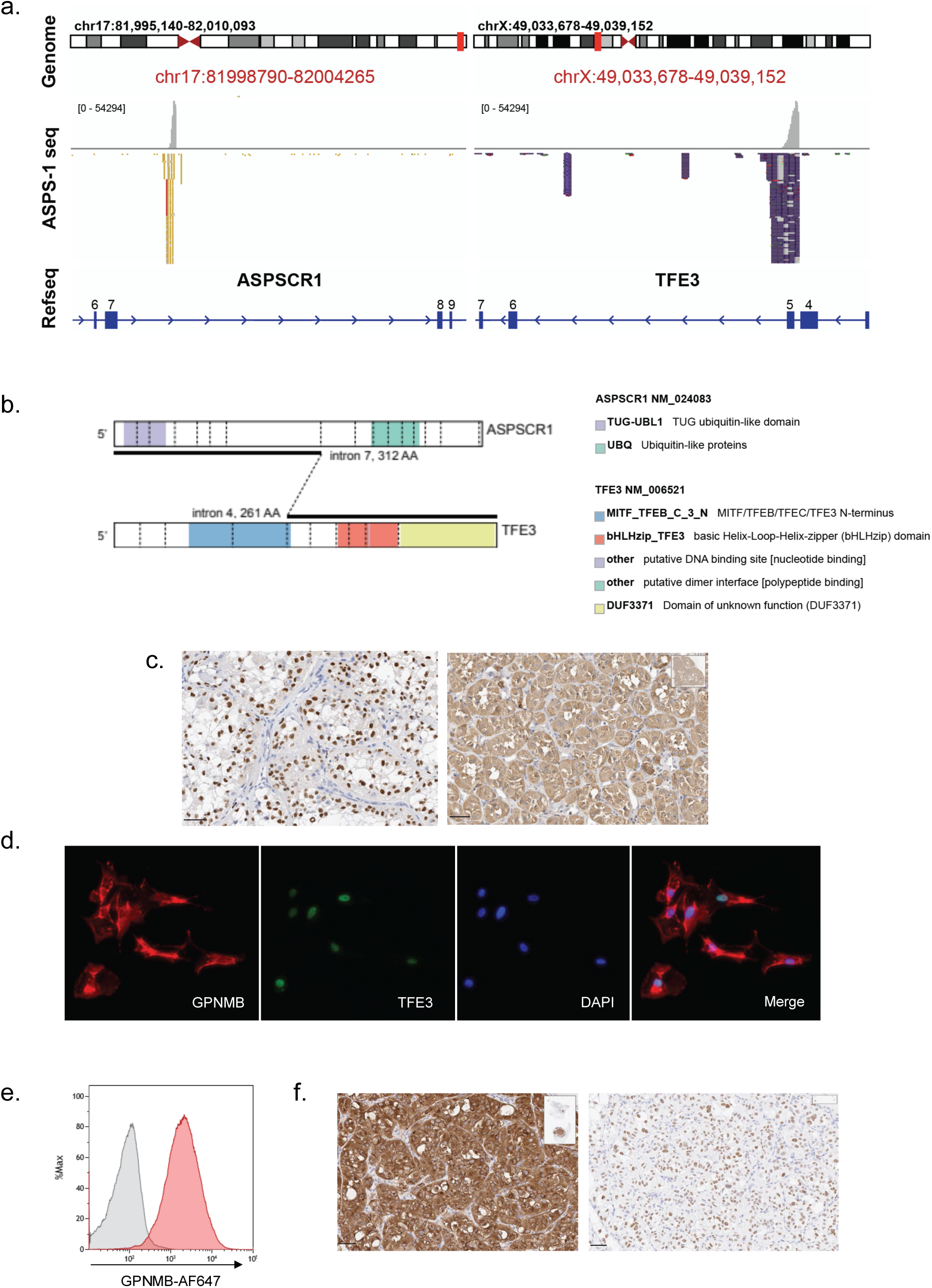
GPNMB Expression in the ASPS PDX-derived Cell Line Model. **a-b,** Amplicon sequencing the patient ASPS-1 demonstrated a type II fusion joining the seventh exon of ASPSCR1 to the fifth exon of TFE3. Sequence alignments of fusion-spanning reads at both gene loci stack at the breakpoint location. Exon numbers are labelled on RefSeq gene models. **c,** TFE3 (left) and GPNMB (right) staining of the primary resection. Error bars = 50 um. **d-e,** Immunofluorescence (d) and flow cytometry (e) of the ASPS PDX-derived cell line. **f,** GPNMB (left) and TFE3 (right) IHC of the ASPS PDX-derived cell line grown intramuscularly in NOD-SCID-MHCKO mice. Scale bars = 50 um.

**Supplementary Figure 4:**
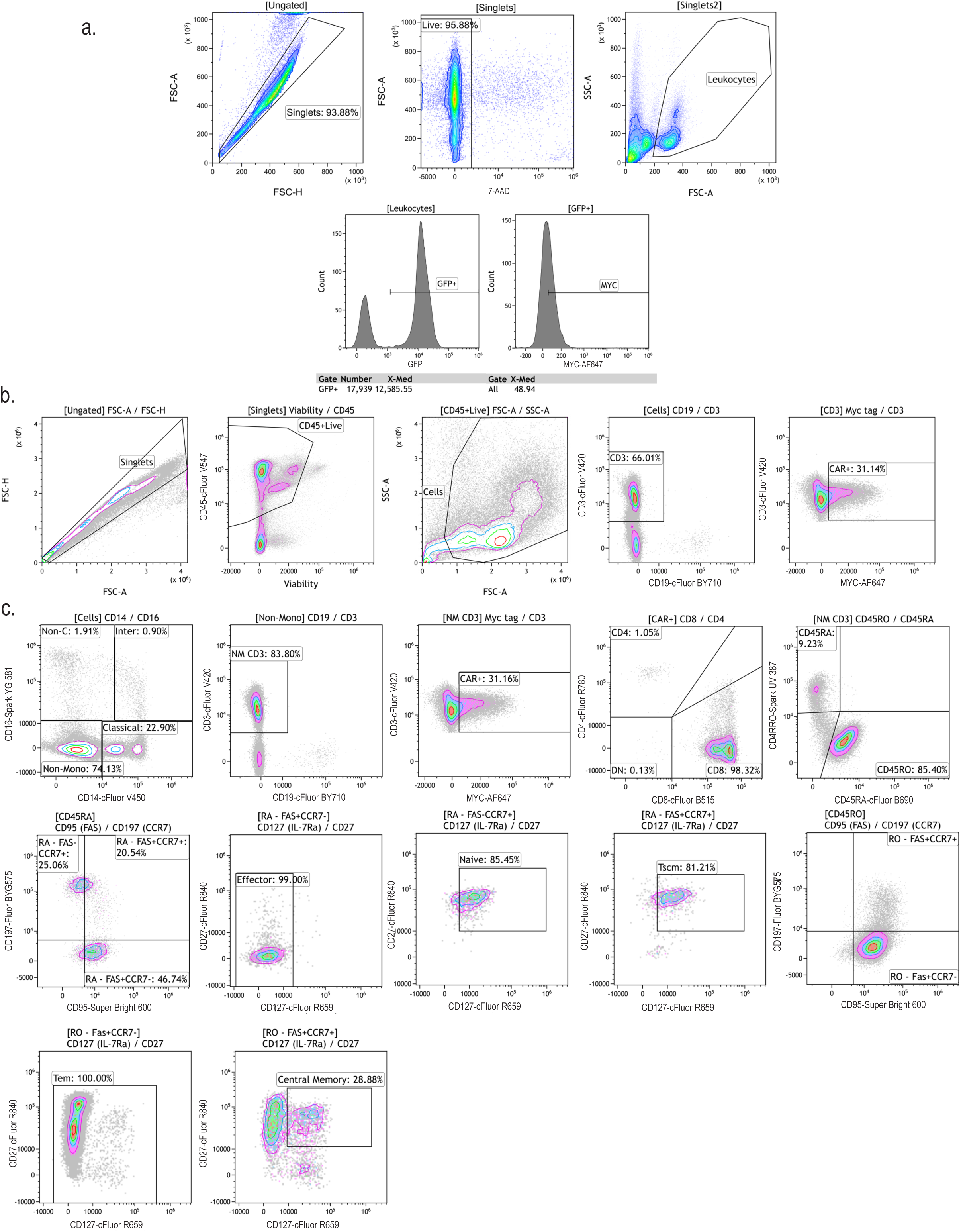
Gating Strategy for Xenograft Bleeds and Clinical Sample. **a,** Representative gating strategy for conventional flow cytometry to enumerate circulating CAR T cells in xenograft models. CAR T cells were counted as GFP+ cells when preclinical vector was utilized. When clinical vector was used, a CD3+MYC+ double gate was employed. **b,** Full spectrum flow gating strategy to enumerate percent CD3+ and CD3+CAR+ cells in blood on Patient ASPS-3. **c**, Full spectrum flow gating strategy to measure circulating GCAR1 memory phenotypes. Naive T cells (Tn; CD45RA+CD45ROCCR7+ CD27+CD127+CD95-), eVector T cells (TeV; CD45RA+CD45RO-CCR7-CD27+/-CD127-CD95+), eVector memory cells (Tem; CD45RA-CD45RO+CCR7-CD27+/-CD127+/-CD95+), central memory T cells (Tcm; CD45RA CD45RO+CCR7+CD27+CD127+ CD95+), stem cell memory T cells (CD45RA+CD45ROCCR7+ CD27+CD127+CD95+) and other (did not fit any of these populations). Cells gated on Live, CD45+CD3+MYC+CD19-CD14-CD16-cells.

**Supplementary Figure 5:**
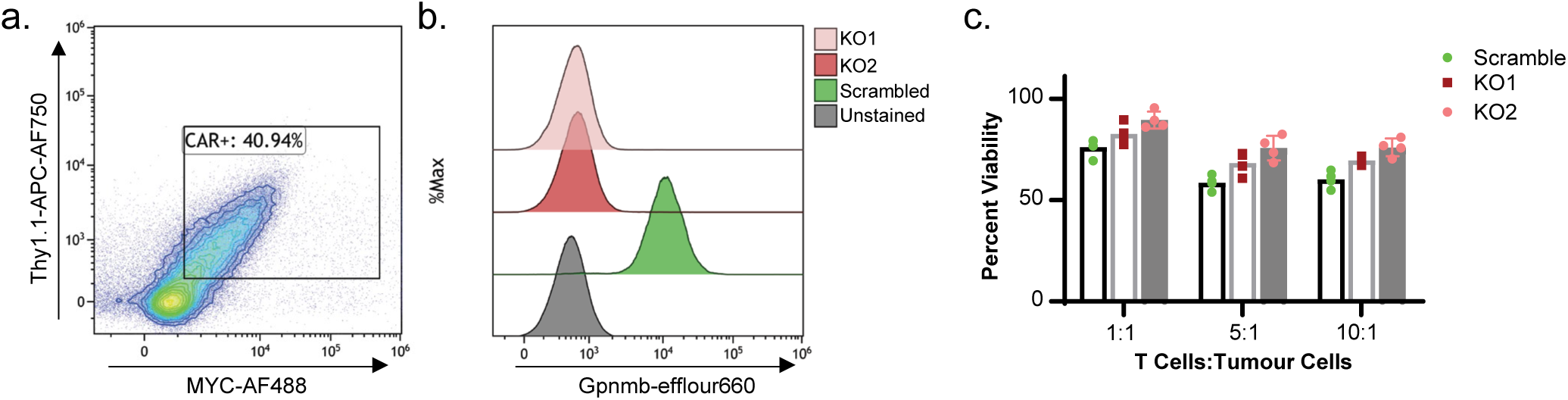
GCAR1 engineered into a mouse CAR vector and tested on B16F10 GPNMB Knockout cells. **a,** Thy1.1 and MYC expression of the G1 scFV from GCAR1 into a mouse a 2nd generation CAR backbone (CD28.z) containing a Thy1.1 transduction marker into C57Bl/6 mouse T cells. **b,** B16F10 mouse cell lines that naturally express Gpnmb (Scrambled) where knocked out for Gpnmb (KO1, KO2). **c,** B16F10 KO and scrambled cell lines were transduced to express an Cherry and firefly luciferase construct, then co-cultured with mouse GPNMB CAR and measured for cytotoxicity 24-hours later with a luminescent assay. Bars represent mean and lines standard deviation of three internal replicates.

**Supplementary Figure 6:**
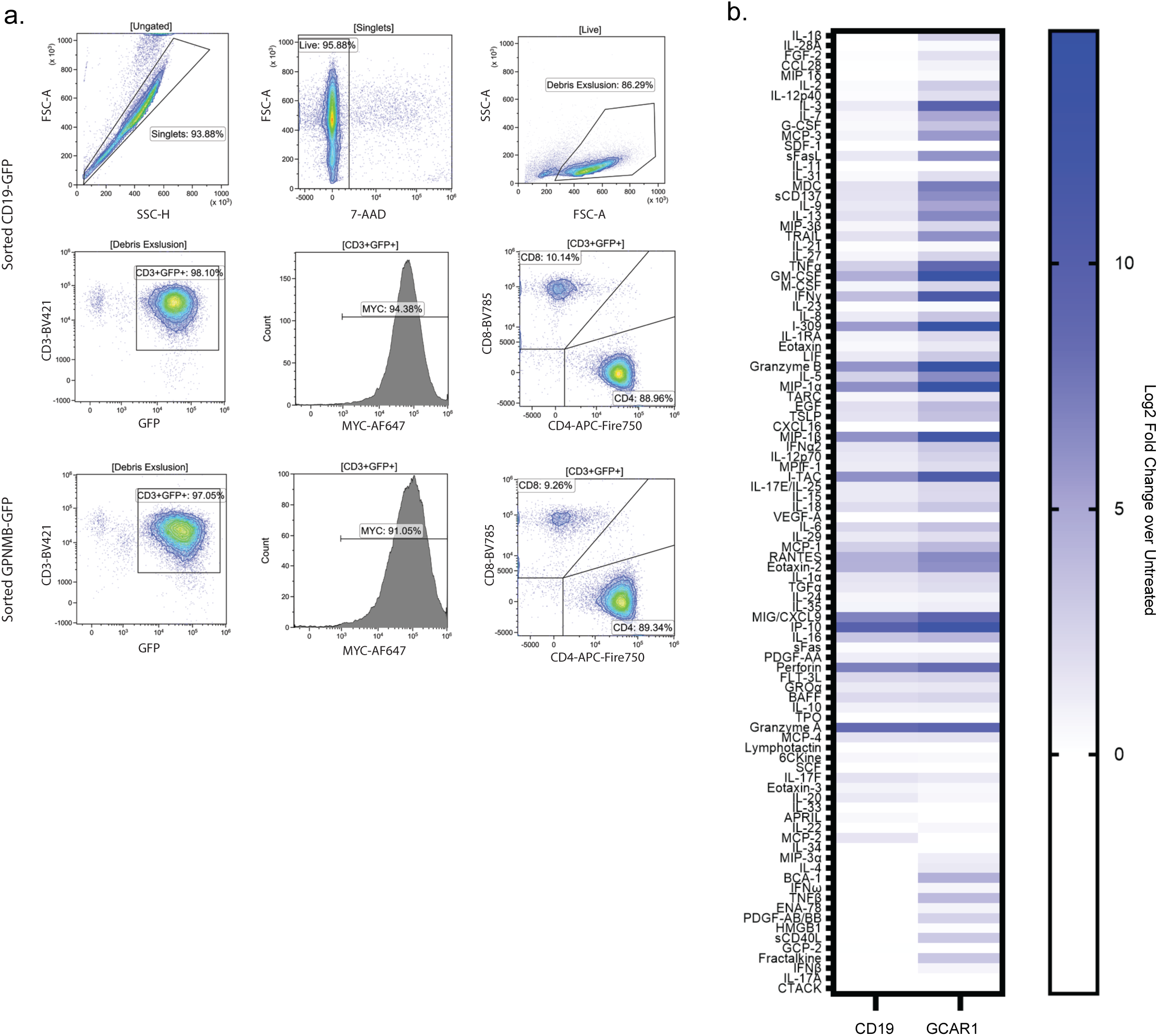
ASPS Patient Explant Preclinical T cell Manufacturing and Cytokine Secretion. **a,** Flow cytometric analysis of preclinically manufactured CD19-GFP of GCAR1-GFP CAR T cells 7 days post transduction from Patient ASPS-2. **b**, Results of 96-plex Luminex assay from media harvested from explants 48 hours after co-culture with 1e4 CAR T cell. Results represent mean of 3 explant cultures from each CAR treatment and displayed as Log2 transformed fold change from untreated media. Results ordered in fold change difference between GPNMB and CD19 CAR.

**Supplementary Figure 7:**
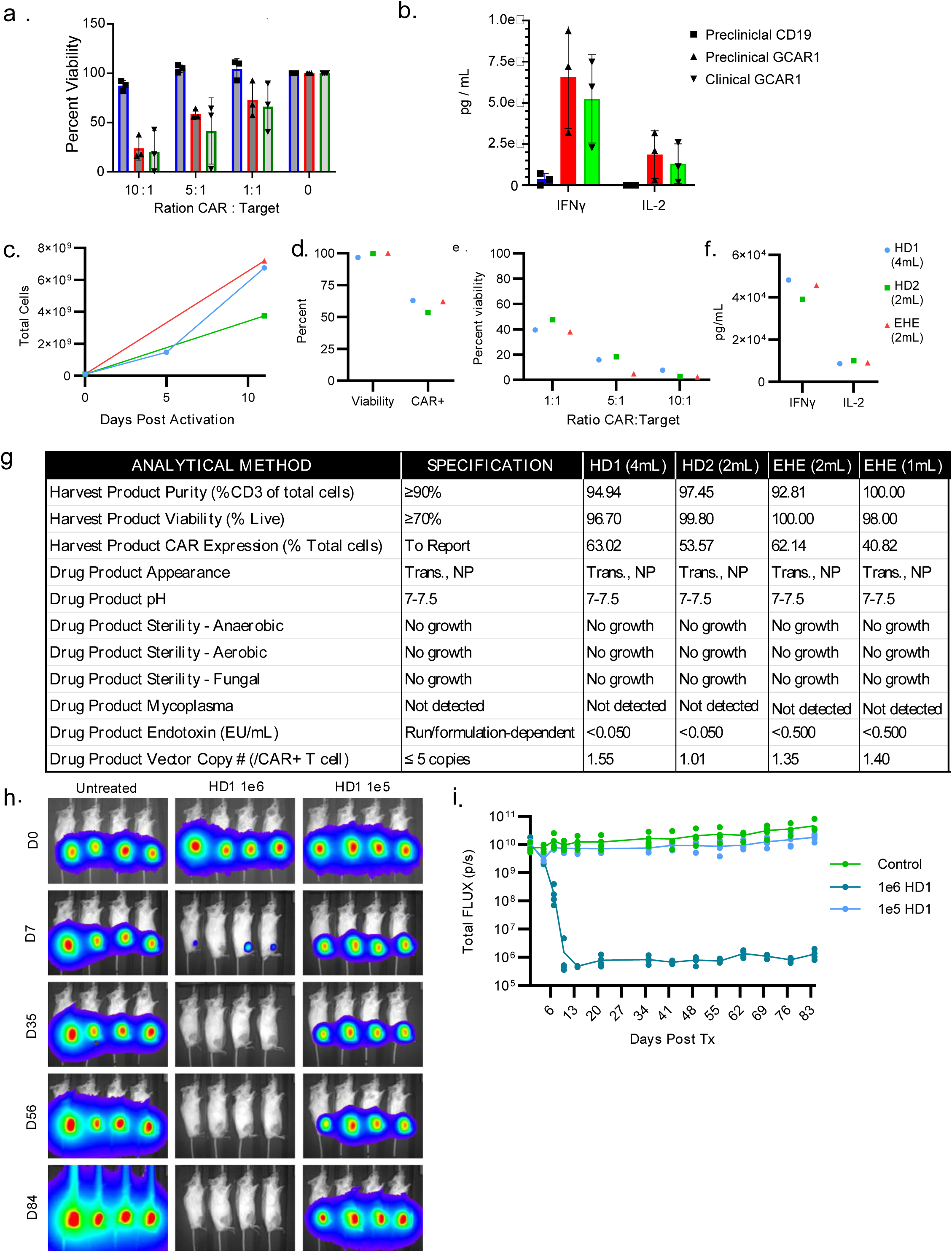
GCAR1 Clinical Production QC Runs. **a-b,** Cytotoxicity (a) and cytokine secretion (b) of laboratory manufactured preclinical or clinical GCAR1 LV vector on ASPS^mc/FLUC^ cells. Graph represent means with standard deviation from n=3 independent repeats in patient ASPS-02 cells. **c-f,** 1e8 magnetically separated CD4 and CD8 T cells were run on a CliniMACS Prodigyc and transduced with indicated volume of GMP manufactured clinical GCAR1 vector. Total T cell expansion (c), viability and CAR transduction efficiency (d), cytotoxicity (e) and cytokine secretion (f) was measured in each run. e, All runs passed qualification standards. h-i, Clinically manufactured GCAR1 behaved similarly in the ASPS PDX-derived model as seen with the preclinical vector (**Extended Figure 2**).

**Supplementary Figure 8:**
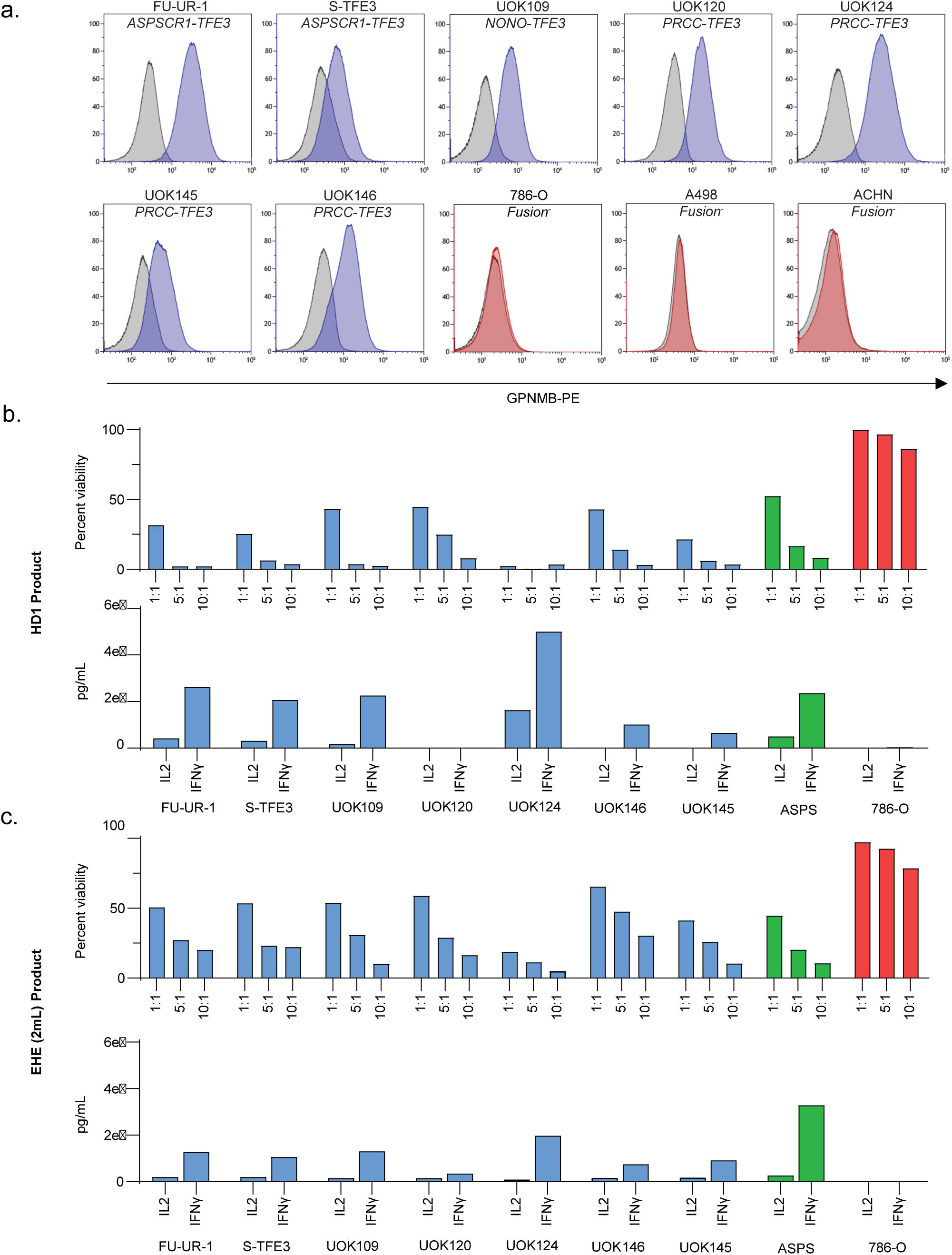
Clinically Manufactured GCAR1 on tRCC Cell Lines in vitro. **a,** Flow cytometric analysis on tRCC cell (blue) or ccRCC cell lines (red) for cell surface expression of GPNMB. **b-c,** All lines were treated with GMP grade, clinically manufactured GCAR1 cells in vitro with the HD1 product (b) or the EHE product (c) and measured for cytotoxicity (top) and cytokine secretion (bottom). All cell lines were transduced with a mCherry and firefly luciferase construct. Cells were co-cultured for 24-hours.

**Supplementary Figure 9:**
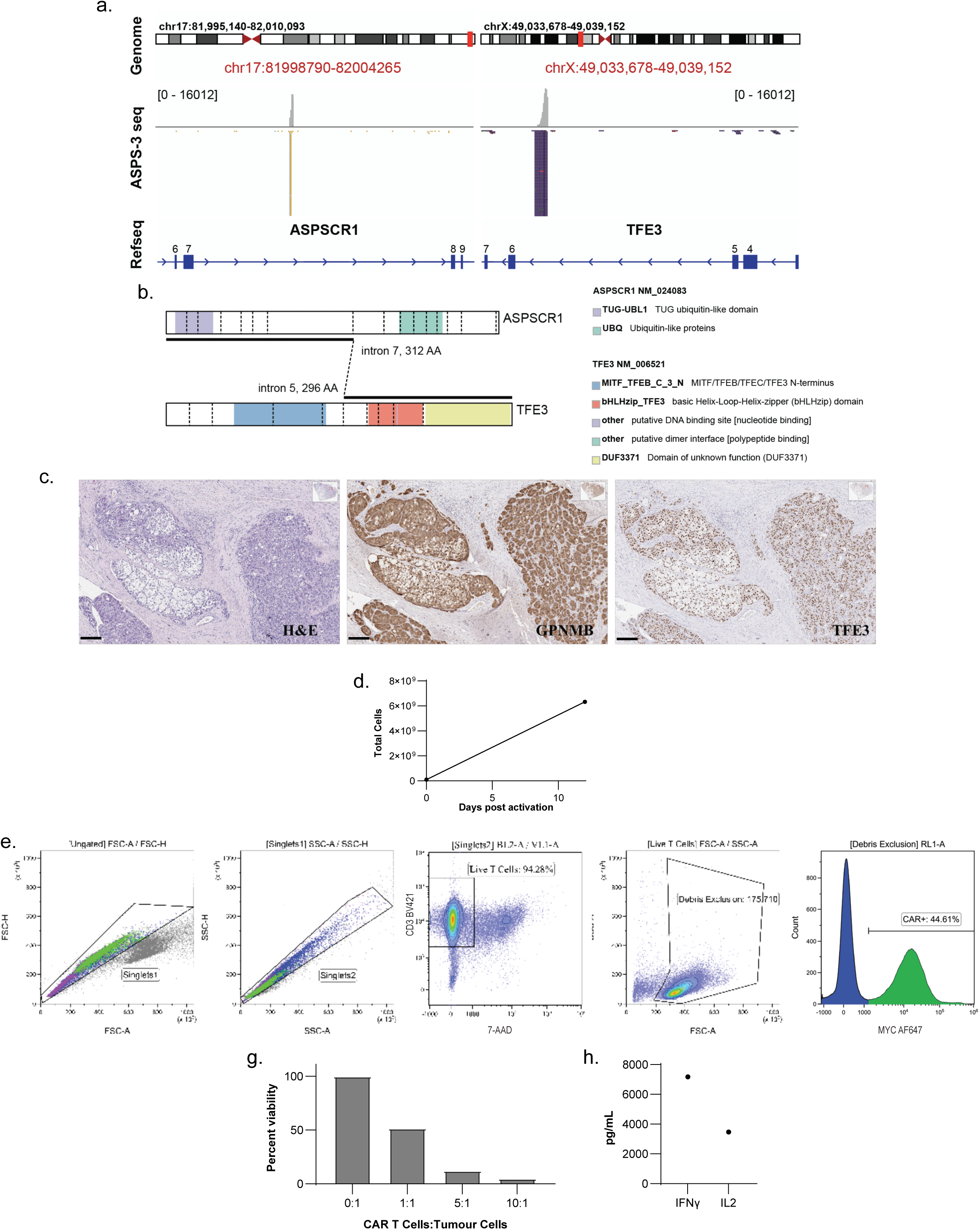
First-in-human ASPS patient primary tumor and clinical GCAR1 product. **a-b,** Amplicon sequencing the patient ASPS-3 demonstrated a type I fusion joining the seventh exon of ASPSCR1 to the sixth exon of TFE3. Sequence alignments of fusion-spanning reads at both gene loci stack at the breakpoint location. Exon numbers are labelled on RefSeq gene models (a). Schematic of the resulting fusion transcript (b). **c,** Immunohistochemical analysis of primary patient tumor resection tissue. **d-f,** 1e8 magnetically separated CD4 and CD8 T cells were run on a CliniMACS Prodigyc and transduced with indicated 2mL of GMP manufactured clinical GCAR1 vector. Total T cell expansion (d), purity and viability, (e) and transduction efficiency (f) measured at 12-day harvest. g-h, Potency was measured by cytotoxic activity of ASPS PDX-derived cell line (g) and cytokine secretion (h).

**Supplementary Figure 10:**
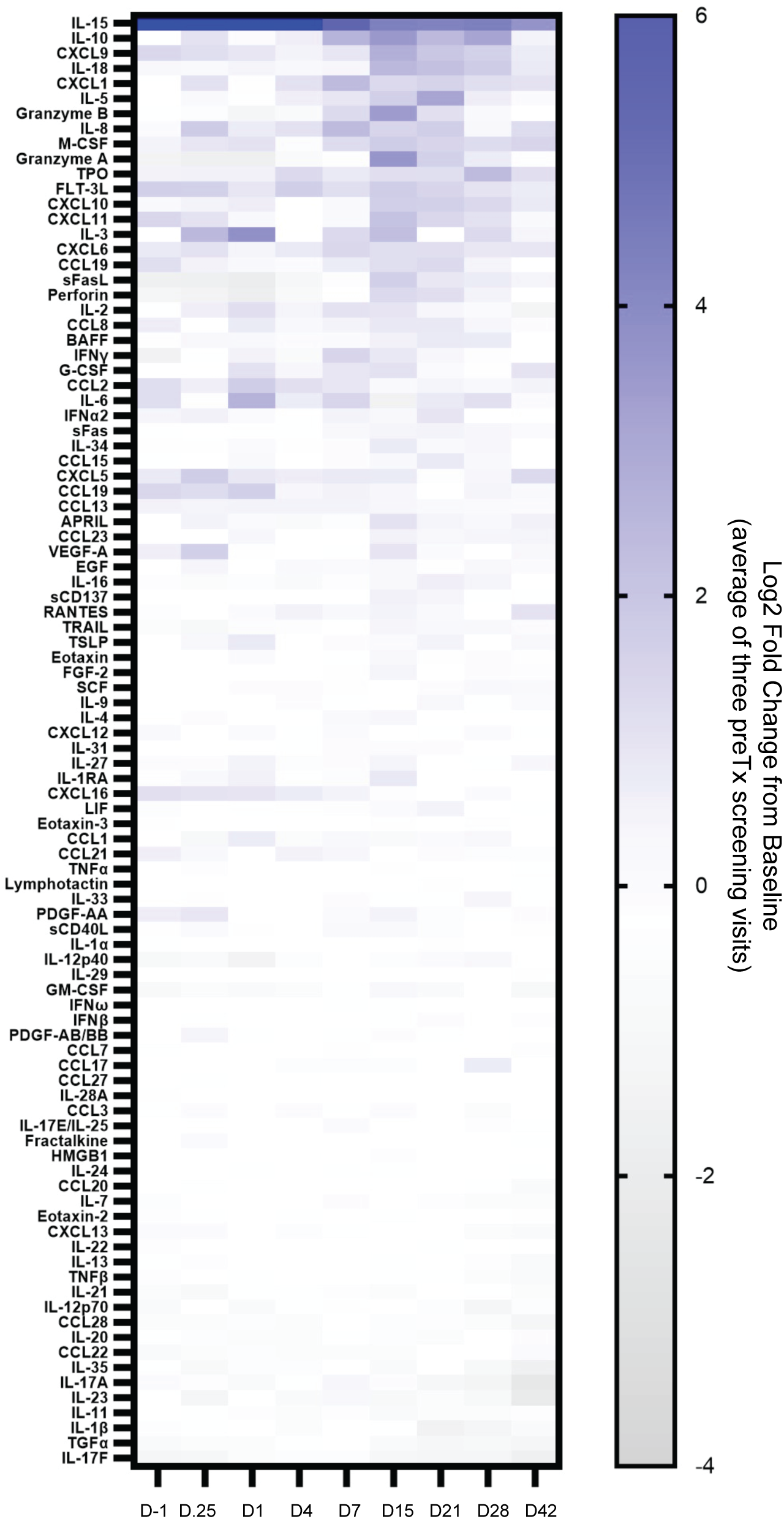
96-plex Luminex of plasma cytokine levels post GCAR1 treatment. Plasma samples drawn from patient at indicated timepoints were measured for analytes using a 96-plex Luminex panel. Results are Log2 transformed fold change from baseline, which was derived from the average of three pre-treatment screening visits.

**Supplementary Figure 11:**
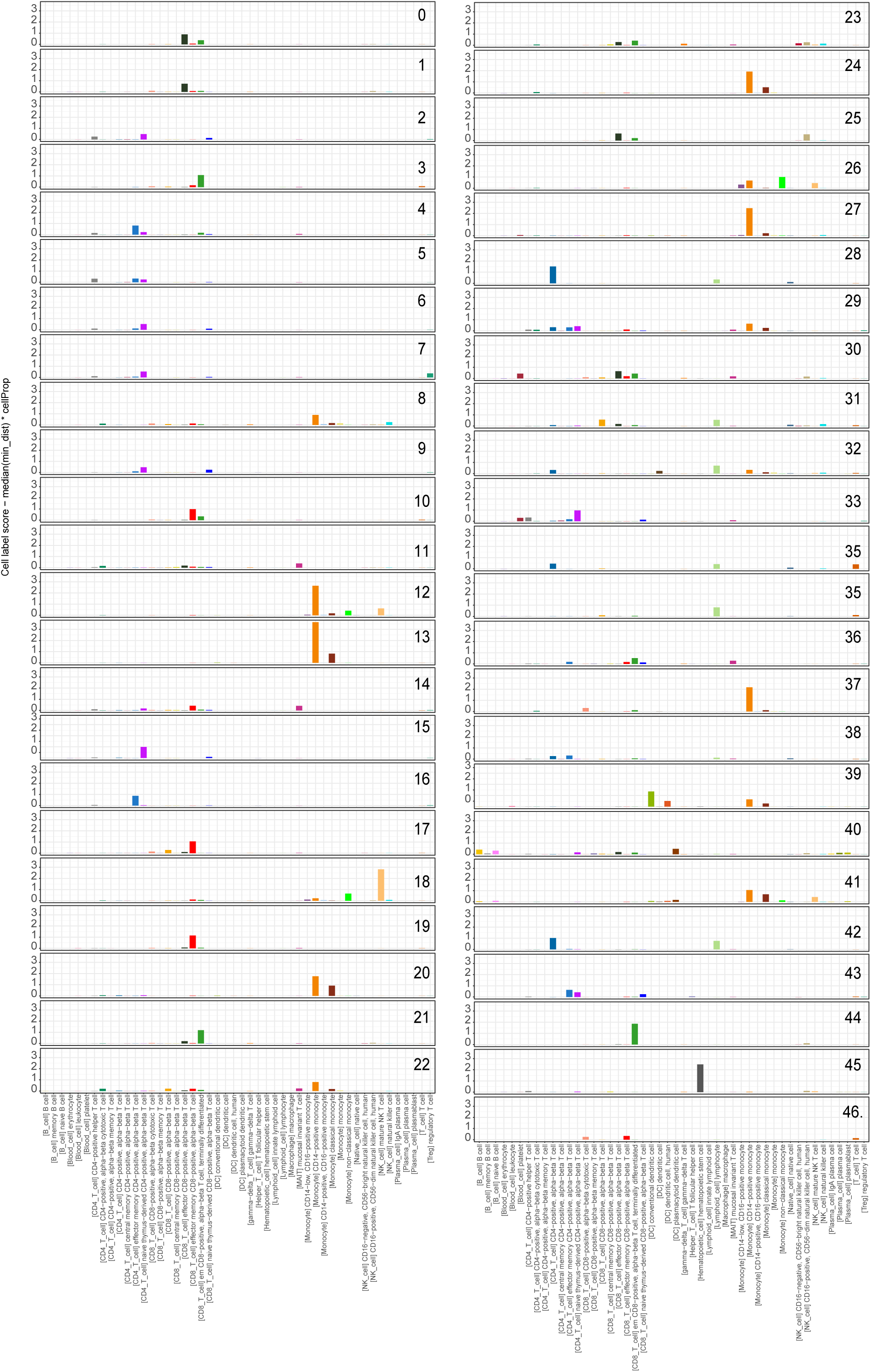
Single cell cluster annotation. Barplots of cell label scores (y-axis) for each cell label (x-axis) are presented per cluster. The score incorporates the SCimilarity min_dist metric (quantifying divergence from reference cell types) and the proportion of cells with each label in a given cluster (see Methods).

## REFERENCES

1. Shah, N. N. et al. Long-Term Follow-Up of CD19-CAR T-Cell Therapy in Children and Young Adults With B-ALL. J Clin Oncol 39, 1650–1659 (2021).

2. Hegde, M. et al. Tumor response and endogenous immune reactivity after administration of HER2 CAR T cells in a child with metastatic rhabdomyosarcoma. Nat Commun 11, 3549 (2020).

3. Brown, C. E. et al. Regression of Glioblastoma after Chimeric Antigen Receptor T-Cell Therapy. N Engl J Med 375, 2561–9 (2016).

4. Del Bufalo, F. et al. GD2-CART01 for Relapsed or Refractory High-Risk Neuroblastoma. N Engl J Med 388, 1284–1295 (2023).

5. Morgan, R. A. et al. Case report of a serious adverse event following the administration of T cells transduced with a chimeric antigen receptor recognizing ERBB2. Mol Ther 18, 843–51 (2010).

6. O’Rourke, D. M. et al. A single dose of peripherally infused EGFRvIII-directed CAR T cells mediates antigen loss and induces adaptive resistance in patients with recurrent glioblastoma. Sci Transl Med 9, (2017).

7. Perry, J., Seong BYA & Stegmiaer. Biology and Therapy of Dominant Fusion Oncoproteins Involving Transcription Factor and Chromatin Regulators in Sarcomas. Annu Rev Cancer Biol 3, 299–321 (2019).

8. Gröbner, S. N. et al. The landscape of genomic alterations across childhood cancers. Nature 555, 321–327 (2018).

9. Xie, L., Zhang, Y. & Wu, C.-L. Microphthalmia family of transcription factors associated renal cell carcinoma. Asian J Urol 6, 312–320 (2019).

10. Bradner, J. E., Hnisz, D. & Young, R. A. Transcriptional Addiction in Cancer. Cell 168, 629–643 (2017).

11. Lazaratos, A.-M., Annis, M. G. & Siegel, P. M. GPNMB: a potent inducer of immunosuppression in cancer. Oncogene 41, 4573–4590 (2022).

12. Maric, G., Rose, A. A., Annis, M. G. & Siegel, P. M. Glycoprotein non-metastatic b (GPNMB): A metastatic mediator and emerging therapeutic target in cancer. Onco Targets Ther 6, 839–52 (2013).

13. Saade, M., Araujo de Souza, G., Scavone, C. & Kinoshita, P. F. The Role of GPNMB in Inflammation. Front Immunol 12, 674739 (2021).

14. Baba, M. et al. TFE3 Xp11.2 Translocation Renal Cell Carcinoma Mouse Model Reveals Novel Therapeutic Targets and Identifies GPNMB as a Diagnostic Marker for Human Disease. Mol Cancer Res 17, 1613–1626 (2019).

15. Kobos, R. et al. Combining integrated genomics and functional genomics to dissect the biology of a cancer-associated, aberrant transcription factor, the ASPSCR1-TFE3 fusion oncoprotein. J Pathol 229, 743–754 (2013).

16. Lee, C.-J. et al. Establishment of an Academic Tissue Microarray Platform as a Tool for Soft Tissue Sarcoma Research. Sarcoma 2021, 6675260 (2021).

17. Salles, D. C. et al. GPNMB expression identifies TSC1/2/mTOR-associated and MiT family translocation-driven renal neoplasms. J Pathol 257, 158–171 (2022).

18. Tse, K. F. et al. CR011, a fully human monoclonal antibody-auristatin E conjugate, for the treatment of melanoma. Clin Cancer Res 12, 1373–82 (2006).

19. Li, W. et al. Chimeric Antigen Receptor Designed to Prevent Ubiquitination and Downregulation Showed Durable Antitumor Efficacy. Immunity 53, 456–470.e6 (2020).

20. Mrass, P. et al. Random migration precedes stable target cell interactions of tumor-infiltrating T cells. J Exp Med 203, 2749–61 (2006).

21. Lau, D. et al. Intravital Imaging of Adoptive T-Cell Morphology, Mobility and Trafficking Following Immune Checkpoint Inhibition in a Mouse Melanoma Model. Front Immunol 11, 1514 (2020).

22. Paoluzzi, L. & Maki, R. G. Diagnosis, Prognosis, and Treatment of Alveolar Soft-Part Sarcoma: A Review. JAMA Oncol 5, 254–260 (2019).

23. Eisenhauer, E. A. et al. New response evaluation criteria in solid tumours: revised RECIST guideline (version 1.1). Eur J Cancer 45, 228–47 (2009).

24. Heimberg, G. et al. Scalable querying of human cell atlases via a foundational model reveals commonalities across fibrosis-associated macrophages. Preprint at 10.1101/2023.07.18.549537 (2023).

25. Conley, A. P. et al. Positive Tumor Response to Combined Checkpoint Inhibitors in a Patient With Refractory Alveolar Soft Part Sarcoma: A Case Report. J Glob Oncol 1–6 (2018) doi:10.1200/JGO.2017.009993.

26. Sterner, R. C. & Sterner, R. M. CAR-T cell therapy: current limitations and potential strategies. Blood Cancer J 11, 69 (2021).

27. Manoharan, V. T. et al. Spatiotemporal modeling reveals high-resolution invasion states in glioblastoma. Genome Biol 25, 264 (2024).

28. Chen, A. P. et al. Atezolizumab for Advanced Alveolar Soft Part Sarcoma. New England Journal of Medicine 389, 911–921 (2023).

29. Biswas, K. B. et al. GPNMB is expressed in human epidermal keratinocytes but disappears in the vitiligo lesional skin. Sci Rep 10, 4930 (2020).

30. Kopp, L. M. et al. Phase II trial of the glycoprotein non-metastatic B-targeted antibody-drug conjugate, glembatumumab vedotin (CDX-011), in recurrent osteosarcoma AOST1521: A report from the Children’s Oncology Group. Eur J Cancer 121, 177–183 (2019).

31. Vahdat, L. T. et al. Glembatumumab vedotin for patients with metastatic, gpNMB overexpressing, triple-negative breast cancer (‘METRIC’): a randomized multicenter study. NPJ Breast Cancer 7, 57 (2021).

32. Yardley, D. A. et al. EMERGE: A Randomized Phase II Study of the Antibody-Drug Conjugate Glembatumumab Vedotin in Advanced Glycoprotein NMB-Expressing Breast Cancer. J Clin Oncol 33, 1609–19 (2015).

33. Bendell, J. et al. Phase I/II study of the antibody-drug conjugate glembatumumab vedotin in patients with locally advanced or metastatic breast cancer. J Clin Oncol 32, 3619–25 (2014).

34. Hasanov, M. et al. A Phase II Study of Glembatumumab Vedotin for Metastatic Uveal Melanoma. Cancers (Basel) 12, (2020).

35. Ott, P. A. et al. Phase I/II study of the antibody-drug conjugate glembatumumab vedotin in patients with advanced melanoma. J Clin Oncol 32, 3659–66 (2014).

36. Ott, P. A. et al. A phase 2 study of glembatumumab vedotin, an antibody-drug conjugate targeting glycoprotein NMB, in patients with advanced melanoma. Cancer 125, 1113–1123 (2019).

37. Bishop, M. R. et al. Second-Line Tisagenlecleucel or Standard Care in Aggressive B-Cell Lymphoma. N Engl J Med 386, 629–639 (2022).

38. Kim, S. I., Cassella, C. R. & Byrne, K. T. Tumor Burden and Immunotherapy: Impact on Immune Infiltration and Therapeutic Outcomes. Front Immunol 11, 629722 (2020).

39. Hanahan, D., Michielin, O. & Pittet, M. J. Convergent inducers and effectors of T cell paralysis in the tumour microenvironment. Nat Rev Cancer (2024) doi:10.1038/s41568-024-00761-z.

40. Huang, Y.-H. et al. Expression pattern and prognostic impact of glycoprotein non-metastatic B (GPNMB) in triple-negative breast cancer. Sci Rep 11, 12171 (2021).

41. Logun, M. et al. Patient-derived glioblastoma organoids as real-time avatars for assessing responses to clinical CAR-T cell therapy. Cancer Stem Cell, ISSN 1934-5909 (2024).

## METHODS REFERENCES

1. Binder, J. X. et al. COMPARTMENTS: unification and visualization of protein subcellular localization evidence. Database (Oxford) 2014, bau012 (2014).

2. Bausch-Fluck, D. et al. A mass spectrometric-derived cell surface protein atlas. PLoS One 10, e0121314 (2015).

3. Uhlén, M., et al. Proteomics. Tissue-based map of the human proteome. Science 347, 1260419 (2015).

4. Bausch-Fluck, D. et al. The in silico human surfaceome. Proc Natl Acad Sci U S A 115, E10988– E10997 (2018).

5. Waas, M. et al. SurfaceGenie: a web-based application for prioritizing cell-type-specific marker candidates. Bioinformatics 36, 3447–3456 (2020).

6. Selvarajah, S. et al. High-resolution array CGH and gene expression profiling of alveolar soft part sarcoma. Clin Cancer Res 20, 1521–30 (2014).

7. Kummar, S. et al. Cediranib for metastatic alveolar soft part sarcoma. J Clin Oncol 31, 2296–302 (2013).

8. Kim, M.-S. et al. A draft map of the human proteome. Nature 509, 575–81 (2014).

9. Wang, D. et al. A deep proteome and transcriptome abundance atlas of 29 healthy human tissues. Mol Syst Biol 15, e8503 (2019).

10. Kuan, C.-T. et al. Affinity-matured anti-glycoprotein NMB recombinant immunotoxins targeting malignant gliomas and melanomas. Int J Cancer 129, 111–21 (2011).

11. Turk, M., Naumenko, V., Mahoney, D. J. & Jenne, C. N. Tracking Cell Recruitment and Behavior within the Tumor Microenvironment Using Advanced Intravital Imaging Approaches. Cells 7, (2018).

12. Beltman, J. B., Marée, A. F. M. & de Boer, R. J. Analysing immune cell migration. Nat Rev Immunol 9, 789–98 (2009).

13. Borcherding, N., Bormann, N. L. & Kraus, G. scRepertoire: An R-based toolkit for single-cell immune receptor analysis. F1000Res 9, 47 (2020).

14. Zheng, G. X. Y. et al. Massively parallel digital transcriptional profiling of single cells. Nat Commun 8, 14049 (2017).

15. Hao, Y. et al. Dictionary learning for integrative, multimodal and scalable single-cell analysis. Nat Biotechnol 42, 293–304 (2024).

16. Scrucca, L., Fraley, C., Murphy, T. B. & Adrian E., R. Model-Based Clustering, Classification, and Density Estimation Using Mclust in R. (Chapman and Hall/CRC, Boca Raton, 2023). doi:10.1201/9781003277965.

17. Heimberg, G. et al. Scalable querying of human cell atlases via a foundational model reveals commonalities across fibrosis-associated macrophages. Preprint at 10.1101/2023.07.18.549537 (2023).

18. Kolberg, L., Raudvere, U., Kuzmin, I., Vilo, J. & Peterson, H. gprofiler2 -- an R package for gene list functional enrichment analysis and namespace conversion toolset g:Profiler. F1000Res 9, 709 (2020).

19. Raudvere, U. et al. g:Profiler: a web server for functional enrichment analysis and conversions of gene lists (2019 update). Nucleic Acids Res 47, W191–W198 (2019).

20. Manoharan, V. T. et al. Spatiotemporal modeling reveals high-resolution invasion states in glioblastoma. Genome Biol 25, 264 (2024).

21. Verhey, T. B. et al. mosaicMPI: a framework for modular data integration across cohorts and - omics modalities. Nucleic Acids Res 52, e53–e53 (2024).

22. Ma, R.-Y., Black, A. & Qian, B.-Z. Macrophage diversity in cancer revisited in the era of single-cell omics. Trends Immunol 43, 546–563 (2022).

23. Lai, Y. et al. Multimodal cell atlas of the ageing human skeletal muscle. Nature 629, 154–164 (2024).

24. Abdulla, S. et al. CZ CELL×GENE Discover: A single-cell data platform for scalable exploration, analysis and modeling of aggregated data. Preprint at 10.1101/2023.10.30.563174 (2023)

